# When the outcome is compositional - a method for conducting compositional response linear mixed models for physical activity, sedentary behaviour and sleep research

**DOI:** 10.1101/2025.06.04.25328944

**Authors:** Aaron Miatke, Ty Stanford, Tim Olds, Francois Fraysse, Carol Maher, Josep Antoni Martin-Fernandez, Dot Dumuid

## Abstract

Time use is compositional in nature because time spent in sleep, sedentary behaviour and physical activity will always sum to 24 h/day meaning any increase in one behaviour will necessarily displace time spent in another behaviour(s). Given the link between time use and health, and its modifiable nature, public health campaigns often aim to change the way people allocate their time. However, relatively few studies have investigated how movement-behaviour composition changes longitudinally (with repeated measures) due to experimental design elements or participant characteristics. This may be because most mixed-model packages that account for the random effects of repeated measures do not natively allow for a multivariate outcome such as movement-behaviour composition. In the current paper we provide a practical framework of how to implement a multivariate response linear mixed model to investigate how movement-behaviour compositions change with repeated measures. In an example we show how time is reallocated in children across the school year.

## 1.0 Introduction

Time spent in daily movement behaviours (sleep, sedentary behaviour (SB), physical activity (PA)) has been linked with many health measures, ranging from adiposity to mental health, cognition and mortality risk ^1^; largely investigated in cross-sectional or observational data. Due to its modifiable nature, intervention and health promotion efforts have long attempted to improve specific aspects of time use, such as increasing time spent in physical activity ^2,3^, reducing sedentary time ^4,5^, or improving sleep ^6^. Most statistical analyses of the effectiveness of such interventions consider only one activity in isolation, even though the constant sum constraint of a 24-h day means that other activity(ies) will necessarily undergo compensatory changes. As all the activities are important to health outcomes, they should all be considered when planning and executing movement behaviour interventions^7^. Understanding shifts in time use resulting from natural temporal (e.g. circadian and circannual) cycles and life transitions are also important considerations. It is imperative to be able to include the full 24-h composition of movement behaviours in the same statistical model when assessing the effectiveness of interventions. The inclusion of all raw 24-h movement behaviour variables (min/day) in statistical models has been problematic due to their constant sum constraint and inherent perfect multi-collinearity which may produce spurious results ^8^. This has been overcome through the application of compositional data analysis (CoDA), whereby 24-h compositions are expressed as logratios prior to their inclusion in statistical models ^9^.

While CoDA is now commonly applied when 24-h movement-behaviour compositions are considered as independent or predictor variables, fewer studies have used CoDA to consider movement behaviour compositions as dependent or response variables ^10^. There is currently no accepted methodology for analysing movement behaviour compositions in a multi-level framework that could be used to, for example, assess whether mean compositions change over time with repeated measurements on individuals, if there are intervention or experimental group effects in compositions over time, or if compositions are associated with sociodemographic or environmental factors while accounting for clustered sampling designs. This paper is structured as follows: in Section 1, we provide background on the rationale for CoDA for 24-h movement behaviours. Section 1.1 introduces the log-ratio methodology and how it is applied within regression modelling, and Section 1.2 describes previous frequentist approaches to analysing compositions as the dependent variables, noting the inability of these methods to include multivariate response variables when the data have a multi-level structure. In Section 2, we present a method for a compositional multivariate-response linear mixed model (CMRLMM). Section 3 provides an example of the application of our CMRLMM to real-world data of Australian school children’s movement behaviour patterns across various time points. In Section 4, we provide a concluding discussion on strengths and weaknesses of the new method, and areas for future development and research.

### 1.1 Brief overview of CoDA and application to 24-h movement behaviours

CoDA is used in behavioural epidemiology to analyse time spent across 24-hour movement behaviours, including sleep, sedentary behaviour (SB), and physical activity (PA). The log-ratio methodology transforms raw compositional data into orthonormal log-ratio (olr) coordinates (also known as isometric log-ratio coordinates [ilr]) before their inclusion in compositional regression models. However, current methods struggle to address a common issue in epidemiological studies: the multi-level structure of movement behaviour data caused by repeated measurements or clustered sampling designs.

Indeed, a composition **u** is a vector in 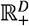 where the only relevant information is contained in the ratios between its components, these components of the composition are aptly named *compositional parts*. For example, the ratio of time awake to time asleep is invariant to whether those times are expressed as proportions of the day, hours or minutes and have specific properties due to the sample space. A D-part composition can be expressed as a vector **x** = [*x*_1_,*x*_2_, …, *x*_*D*_], where all parts are non-negative (but ideally strictly positive) and sum to a positive constant *k*. Typically, its sample space is mathematically written as 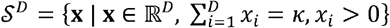 where *S*^*D*^ is known as the *D*-part simplex or *D*-simplex, a (*D* −1) dimensional subset of real space *R*^*D*^ ^11^. Because compositions only contain relative information, a composition **u** can be closed to any positive constant such that

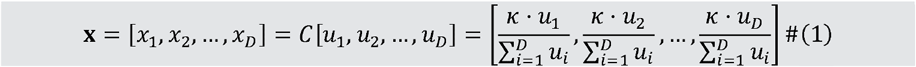

where *C* is the closure operation to the constant *k*. For example, if the parts of the vector represent time spent in different behaviours, the composition could equivalently be closed to *κ* = 1440 (min/day), *κ* =24 (hour/day), *κ* =1 (proportion of day). When considering differences, or changes, in compositions, traditional algebraic addition and subtraction operations for Euclidean space, are not suitable as they are not scale invariant nor sub-compositionally coherent ^12,13^. However, in the simplex space compositions can be perturbed such that **x** ⊕ **y** = 𝒞[*x*_1_*y*_1_,*x*_2_*y*_2_, …, *x*_*D*_*y*_*D*_] which is analogous to addition in real space. In addition, the powering of **x** ∈ *S*^*D*^ by a constant 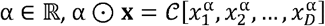, which plays the typical role of the product of a vector by a scalar. Using these operations a linear regression model with compositional response can be formulated as

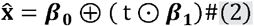

where the coefficients and **β**_0_, **β**_1_ ∈ *S*^*D*^ *t* is a real covariate. Further, the neutral element **1** = 𝒞 [1, …, 1] = [*k*/*D, k*/*D*, … *k*/*D*] has the expected properties such that **x** ⊕ 𝒞 [1, …, 1] = 𝒞 [1, …, 1] ⊕ **x** = **x**. Finally the inverse element of, is defined as*C* [1/ *x*_1_, 1/*x*_2_, … 1/ *x*_D_], denoted as ⊖ **x**. It can be shown that when a composition is perturbed by its inverse it will result in the neutral element such that **x** ⊕ (⊖ **x**) = (⊖ **x**) ⊕ **x** = 𝒞 [1, ⊖,1]. While ‘stay in the simplex’ algebra like that outlined above can be used to summarise or interpret CoDa, most multivariate statistical techniques are not suitable for raw CoDa and require expressing the raw data in terms of log-ratios of the parts. As such, most compositional techniques used in time-use epidemiology favour using the log-ratio approach, specifically the olr transformation ^9^. While other log-ratio approaches have been proposed, such as the additive log-ratio (alr) or centred log-ratio (clr), these have drawbacks that limit their use within statistical models such as mixed models. Namely, alr coordinates are asymmetric, meaning that distances are not preserved within the simplex space. This means results depend on the compositional part chosen as the denominator when constructing the coordinates. Unlike alr coordinates, clr coordinates are isometric, meaning distances are preserved. However, the clr coordinates are also restrictive and spurious when used within statistical models because of the zero-sum constraint ^9,14,15^. Neither of these issues exist when using the olr transformation which is generally preferred in most modelling applications, including within time-use epidemiology ^9^. The olr transformation The olr transformation involves expressing D-part compositional vector of movement behaviour data that exist in the simplex, **x** = [*x*_1_,*x*_2_, …, *x*_*D*_] ∈ *S*^*D*^, into D-1 olr-coordinates ^16,17^ that exist in real space, **z** = olr (**x**) = [*z*_1_,*z*_2_, …, *z*_*D*−1_] ∈ ℝ _*D* − 1_. That is, the vector **z** are the coordinates of composition **x** in an olr-basis. Importantly, these olr-coordinates can be used in standard multivariate statistical models ^18^.For example, the linear regression model in Eq. (2) is expressed in olr-coordinates as olr 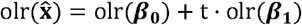.

An olr-basis can be created using a data-driven method such as Principal Balances ^19^ or R-mode cluster analysis ^20^. Alternatively, the knowledge of the researcher can be used to improve the interpretation of the models when creating the olr-basis by a sequential binary partition (SBP) process, which is generally preferred in time-use research (15). An SBP process uses a (D – 1) x D dimensional sign matrix to iteratively divide the compositional parts until all groups consist of a single component ^21^.

The olr-coordinates of the general form can be defined as

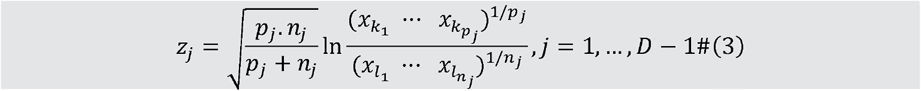

where the *p*_*j*_ and n _*j*_ are the number of parts in the *j-*th row of the sign matrix that are coded positive and negative, respectively and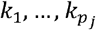 are the indexes of parts in the numerator and 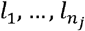 are the indexes of parts in the denominator of each row of the sign matrix ^21^.

Note that the olr-coordinate in Equation (3) is a “balance” between the average of two sets of parts. Balance coordinates may be of use when a researcher is interested in distinct groups of behaviours ^22^ (e.g., active vs. passive behaviours). If a researcher has a particular interest in one behaviour, they may use pivot coordinates of the general form

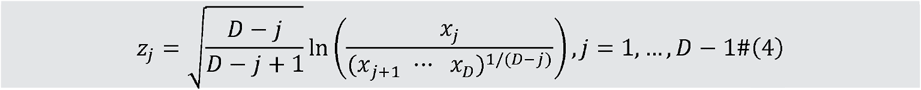

Here, the first olr-coordinate (*z*_1_) reflects the dominance of one behaviour (x_1_) relative to the geometric mean of the remaining behaviours and is of use when a researcher has a particular interest in one behaviour. Note that 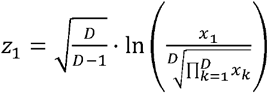 that is, the first olr-coordinate is proportional to the first clr-coordinate The remaining olr-coordinates, *z*_2_, …, *z*_*D* − 1_, are then created in a similar manner with the denominator of the previous coordinate split with until no parts remain^23^.

While hypothesis-driven construction of olr-coordinates may provide one way of interpreting findings from compositional models, there are limitations to how meaningful these contrasts are to everyday life. For example, the first pivot coordinate (*z*_1_) must be interpreted as the relative increase of one activity, while the geometric mean of the remaining activities is reduced. In real life, changes in the geometric mean of a group of activities may be difficult to conceptualise, it is often easier to interpret findings within the simplex space ^24^.

Importantly, while the interpretation for individual olr-coordinates will change depending on the basis chosen when creating the olr-coordinates, collectively they are equivalent and will retain all relative information about the movement behaviour composition no matter how they are constructed such that x = olr ^-1^(**z**) for any olr-basis. This means in order to make inferences about the movement behaviour composition as a whole, the vector of olr-coordinates x = [*z*_1_, *z*_2_, …, *z*_*D*−1_]must be considered collectively. One option is to use the statistical model to compute point estimates for scenarios of interest, relevant to the research question. For example, if the logratios are considered as predictors of a health outcome in a linear regression model, the model parameters can be used to estimate the value of the health outcome for a selection of different movement behaviour compositions that emulate the reallocation of time between activities ^25^. If the logratios are considered as the dependent variables in a multivariate response linear regression model, the model parameters can be used to estimate what the logratios (and the corresponding 24-h movement-behaviour compositions) would be at different levels/values of the predictor or independent variable. However, when considering compositional responses, standard multivariate response linear regression models are often unsuitable in epidemiological studies due to non-independence of observations resulting from repeated measurements on sampling units (e.g., participants; or clustered sampling designs, e.g., participants within health centres). In these instances, a linear mixed-effects model (LMM) that extends the general linear model to include both fixed and random effects that account for correlated observations is a popular and understood method. Importantly, commonly used statistical software packages for LMMs within a frequentist framework (STATA, SPSS, SAS, R) do not support the inclusion of multivariate outcomes natively (without some data manipulation and model re-specification), such as movement behaviour composition expressed as logratios ^26-28^. This difficulty explains why an approach previously, as described in Section 1.2, to investigate longitudinal changes in movement-behaviour composition has been to use multiple univariate response LMMs, each with a different olr-coordinate as the outcome ^29-32^.

### 1.2 Compositional univariate-response linear mixed model

Consider a univariate compositional LMM to investigate changes in movement-behaviour compositions, with repeated measurements on the same individuals (i=1,2,…,N) over T timepoints *ti*_1_, *ti*_2_, …, *ti*_T_. This model will have time as the level 1 response which is nested within individuals as the level 2 response and has been used by some researchers in the general form

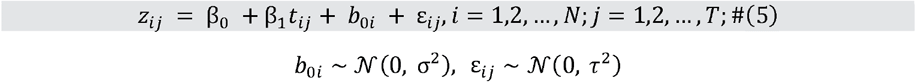

Where *z*_*ij*_ is the response for an individual olr-coordinate, for the *i-*th subject at the *j-*th timepoint, β_0_ is the mean value for when all predictors are equal to zero, is the random b_0*i*_ is the random intercept for *i-*th subject β_1_ is the slope coefficient representing the mean linear slope change over time, timepoint over time, timepoint *t*_*ij*_ is the specific *j*-th time for subject *i*, and ε_*ij*_ is the residual error Previously, researchers have generally investigated compositional outcomes within a multilevel framework using multiple models of the general form Equation (5) in one of two ways. Approach 1) has involved iteratively constructing D sets of pivot coordinates (Equation (4)), with *z*_1_ reflecting the dominance of a different behaviour in each instance and using this coordinate as the dependent variable in D different LMM analyses ^29,30^. Alternatively, approach 2) similarly involves a ‘multiple models’ approach for each component in the vector of olr-coordinates constructed using a single basis ^31,32^, that is, a separate model for each element of the vector **z** = [*z*_1_, *z*_2_, …, *z*_*D*−1_] (Equation (3)).

These methods can be useful approaches; however, they also have limitations. Firstly, both approaches may have an inflated type 1 error rate associated with running repeated analyses with the same data ^33^. Moreover, while approach 1 may be useful if a researcher has a particular interest in a single behaviour, as described earlier the coefficients for a single pivot coordinate are often practically less meaningful to interpret 34. Similar difficulties exist when interpreting results for individual olr coordinates using approach 2, particularly for coordinates *z*_2_, …, [*z*_*D*−1_], where only some behaviours will be involved in their creation. Approach 2 may allow for results to be transformed back into the simplex to allow for more meaningful interpretations via model-based estimates. However, this requires estimates from multiple, independent models to be pieced together. Importantly, approach 2 ignores any correlation structure among the olr-variables. By using *D* − 1 individual models of form Equation (5), we are assuming that the random effects and errors come from separate, and unrelated, normal distributions. However, this is unlikely to be true in the case of CoDa, where the olr-coordinates are intrinsically multivariate in nature and are usually correlated. This means that once transformed back to the simplex, model-based estimates and residuals may differ depending on the sign matrix and associated olr-basis used to construct the coordinates (see examples in supplementary material 1). This is a problem when working with compositional data, where invariance under change of olr basis is one of the fundamental principles ^18^. Indeed, the use of multiple, independent models as outlined in approach 2 above implicitly assumes no relationships between olr coordinates. This is equivalent to specifying a diagonal matrix (heterogenous variance with zero correlation between elements) in the multivariate sense. However, if, after the change of basis, the model is solved again using the multiple-univariate model procedure outlined in approach 2, it is implicitly assumed that the matrices are diagonal, and therefore, the same result as before the change of basis cannot be obtained. Simply put, the only way to obtain equivalent results before and after the change of basis is by using a multivariate procedure that accounts for the relationship between olr coordinates (see supplementary file for further details).

Moreover, while it is possible to test the effects on individual olr-coordinates using the multiple-models approach, there is difficulty in performing a single test of the joint effects on the complete vector of olr-coordinates ^35^, for example, to explore whether the composition of 24-h movement behaviours changed differently between intervention and control groups. In order to overcome these limitations a CMRLMM is required, as presented in Section 2.

## 2. Compositional multivariate-response linear mixed model

We propose extending the univariate mixed model above in Equation (5) to the multivariate case where response vectors are **not** modelled with implied independence. By taking the univariate equation above and adding subscript r to indicate the specific olr-response in question (i.e,*r* = 1,2, *D* − 1] the dependence between response vectors can be more easily specified. Our multivariate formula now becomes:

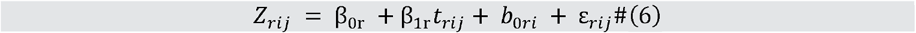

where *Z*_*rij*_ is response *r* for the *i* th participant at the th timepoint; *β*_0*r*_ is the fixed intercept specific to response olr-coordinate; *β*_1*r*_ is the fixed ‘slope’/contrast between time-point *j* and time-point specific to response olr-coordinate Note, the general formula outlined here assumes a linear change over timepoints and is used for simplicity in notation. In instances where this is not true, *β*_1*r*_ *t*_*rij*_ can be replaced to estimate olr-responses over the T timepoints via an indicator function as follows 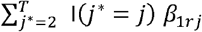 where I(*a*) is an indicator function that is equal to 1 when the argument *a* is true, 0 otherwise So far, this appears similar to the previous formula. However, unlike Equation (5) in Equation (6), the random effects and error will be of length *D* − 1, and are now assumed to come from a single multivariate normal distribution, rather than multiple unrelated univariate distributions, as follows:

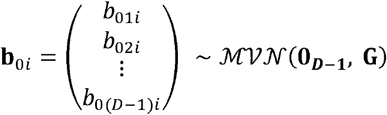

where *b*_0*ri*_are potentially differently varying and correlated random intercepts for each response olr-coordinate *r*, specific to person*i*; and

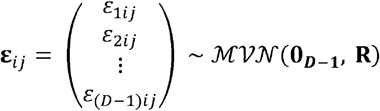

where *ε*_*rij*_ are potentially differently varying and correlated random errors for each response olr-coordinate *r*, specific to person *i* at time-point *j* for each response olr-coordinate *r*; with group- and residual-side variance and covariance (*D* − 1) × (*D* − 1) matrices

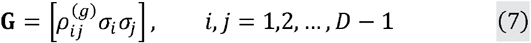

and

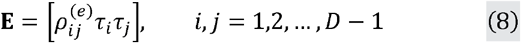

Respectively. Or, written more fully in expanded form

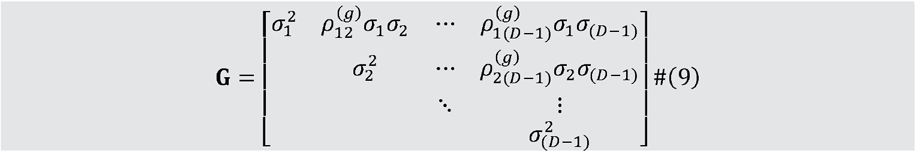

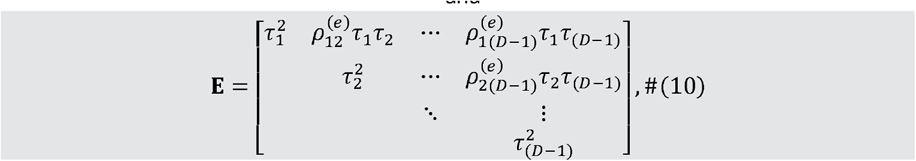

where correlations ρ at the differing levels of model are differentiated with the (*g*) and (*e*) superscripts, where 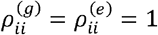. Note that both matrices are symmetrical, that is, **G**_*ij*_= **G**_*ji*_and **E**ij = **E**_*ji*_for any (ij) entry.

Unlike in Equation (5), this model accounts for the correlation between each olr-coordinate by estimating covariances between random effect and residual error terms as shown in the off-diagonal elements above which are constrained to be equal to zero when using the multiple models approach outlined earlier. Thus, the model outlined in Equation (6) allows olr-responses to flexibly vary (and covary) for each olr at both the group (participant) level and residual level. Estimates can then be made for olr-coordinates while respecting the multivariate nature of the data. Just like in the model outlined in Equation (5) the estimates at all levels of the model, including the residuals, will be specific to the basis chosen when constructing the olr-coordinates. However, when (uniquely) transformed back in the simplex space using the inverse transformation orl ^−1^(**β**), the compositional coefficients, and any time-use estimates made using the model, are invariant to the basis used to construct the original olr-coordinates. Currently, in a frequentist framework, the multivariate response linear mixed model of Equation (6) cannot be fitted in R in the most popular linear mixed model packages `nlme` ^36^ and `lme4` ^37^. However, there are two potential solutions to this problem: (a) use a Bayesian framework to fit the multivariate response linear mixed model. For example, using `brms::bms()`; or (b) re-define the data structure to fit an equivalently specified (frequentist) univariate response linear mixed model using `nlme::lme()`and `lme4::lmer()`. The latter is what we pursue in this paper in the hope to create a template for future compositional response linear multilevel models for repeated observations or clustered data as the frequentist framework (knowingly or unknowingly) is probably the most familiar statistical approach of most time-use and other CoDa researchers (although choosing one approach should not be mutually exclusive of the other).

In order to be able to fit Equation (6) using the frequentist framework, the data have to be restructured so that all olr-coordinate responses are in a single column to be used as the dependent variable. We term the re-defined data structuring to fit an equivalently specified univariate response linear mixed model the “stacked response” linear mixed model approach. This terminology is borrowed from the limited online resources ^38,39^, and even fewer published descriptions ^40,41^, and of course how the multivariate response vectors are stacked to make a univariate response vector. Additional dummy variables are then needed to specify the olr-coordinate response in question, that is, a dummy variable for each response r = 1,2,…,D-1 Our original dataset which was of length *ij* (*i* individuals over *j* timepoints each), now becomes length *ijr* with one row per olr-coordinate, per participant, per timepoint. We can then specify a single model that allows for individual changes in each olr-response, while accounting the structure of the random effects and error terms as specified in Equations (9) and (10). An example of the stacking process is outlined below for *r* = 1,2,3 in Tables 1a and 1b. In the restructured dataset the vector of olr-coordinates is contained in a single column along with the dummy variables The dummy variables are binary variables representing which olr-response is being considered. By expanding Equation (5) we can now fit Equation (8) where the response is the notionally ‘univariate’ response associated with the stacked vectors of responses

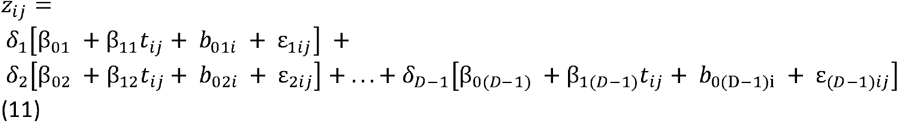

**Table 1a.**
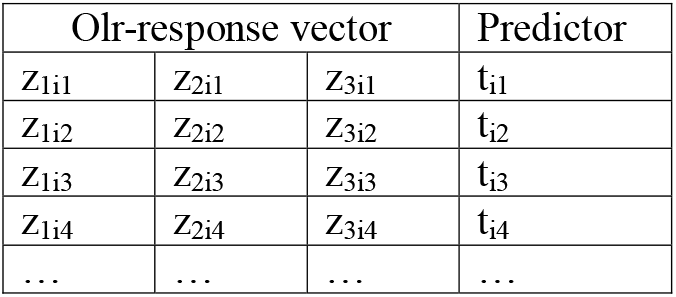
The original data in general form in wide format for the *i*th individual

**Table 1b.** The restructured dataset in general form in long format for the *i*th individual after ‘stacking’ the multivariate olr-response variable.

| Olr value | olr1/Response 1 indicator dummy variable ($\delta_1$) | olr2/Response 2 indicator dummy variable ($\delta_2$) | olr3/Response 3 indicator dummy variable ($\delta_3$) | predictors |
| --- | --- | --- | --- | --- |
| $Z_{1i1}$ | 1 | 0 | 0 | $t_{i1}$ |
| $Z_{2i1}$ | 0 | 1 | 0 | $t_{i1}$ |
| $Z_{3i1}$ | 0 | 0 | 1 | $t_{i1}$ |
| $Z_{1i2}$ | 1 | 0 | 0 | $t_{i2}$ |
| $Z_{2i2}$ | 0 | 1 | 0 | $t_{i2}$ |
| $Z_{3i2}$ | 0 | 0 | 1 | $t_{i2}$ |
| $Z_{1i3}$ | 1 | 0 | 0 | $t_{i3}$ |
| $Z_{2i3}$ | 0 | 1 | 0 | $t_{i3}$ |
| $Z_{3i3}$ | 0 | 0 | 1 | $t_{i3}$ |
| $Z_{1i4}$ | 1 | 0 | 0 | $t_{i4}$ |
| $Z_{2i4}$ | 0 | 1 | 0 | $t_{i4}$ |
| $Z_{3i4}$ | 0 | 0 | 1 | $t_{i4}$ |
| ... | ... | ... | ... | ... |

Or more succinctly expressed as

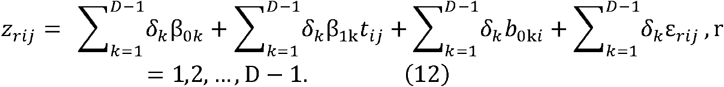

Where *δ*_1_, *δ*_2_, …, *δ*_*D* − 1_, are the olr (or response index) indicator variables associated with each olr-response as specified in Table 1b where the value of *δ*_*r*_ = 1 for the rows of data containing the dependent variable *r*, and 0 otherwise. Thus, the dummy variables act as an additional level to the model which simply defines the response structure. Of note, when compared to the univariate mixed model in Equation (5), the multivariate model in this form also no longer has an overall intercept. Instead, the intercept for each olr-response is estimated separately based on the dummy coding used. Equation (11) can now be used to estimate the vector of responses simultaneously while respecting the multivariate nature of the data. Expanding the formula to include additional demographic predictors such as socio-economic status, body-mass index etc. is then relatively simple. These can be included in the model call by including interaction terms between the dummy coding used and variables of interest to allow each fixed effect to have a specific response (index) associated estimate.

### Multivariate test on fixed effects

Another key advantage of the CMRLMM is the ability to perform a multivariate test on the fixed effects, such as a multivariate F test or Wald chi-square test to test for significance in changes in composition across timepoints, between groups, or other variables of interest. When using the multiple models approach outlined in Section 1.2, it is only possible to perform univariate tests on individual olr-coordinates. However, when using a CMRLMM, it is also possible to conduct tests that are multivariate in nature because the model includes the complete vector of olr-coordinates **z**_*ij*_ = [*z*_1*ij*_, *z*_2*ij*_, …, (*D* −1)_*ij*_] Moreover, because the covariances between are accounted for at both levels of the model, the compositional representation of the model is equivalent regardless of the basis chosen when constructing the olr-coordinates, meaning results will be consistent.

## 3. Example

In this section we use data from the Life on Holidays (LoH) study, for which a full protocol describing data collection methods has been published previously ^42^. LoH was a longitudinal cohort study based in Adelaide, Australia, that aimed to track changes in 24-h activity composition, diet and weight status of primary school-aged children during the school year and summer school holiday periods (n=241). Time use was measured using wrist-worn GENEActiv accelerometers at five timepoints across two school years between 2019-2021: Timepoint 1, at the start of Grade 4 (February-March); Timepoint 2, at the end of Grade 4 (October-November); Timepoint 3, during the summer holiday period; Timepoints 4 & 5, at the start and end of the Grade 5 school year, respectively. Time-use composition was conceptualised as a 4-part composition (D= 4) consisting of time spent in sleep, sedentary behaviour (SB), light PA (LPA) or moderate-to-vigorous PA (MVPA). Each minute of the day was subsequently classified as either SB, LPA or MVPA using validated cutpoints ^43^ with sleep time distinguished from waking time using the algorithm published by Van Hees, Sabia ^44^. Unconditional CMRLMM was initially created without the addition of any covariates. Baseline categorical socio-economic status as determined by parental income, and continuous BMI z-score were then included in the full model as an example of how to include time-invariant covariates.

Children’s movement-behaviour compositions were expressed as olr-coordinates using the SBP and sign matrix shown below, where the 4-part compositional vector**x** = [*x*_1_, *x*_2_, *x*_3_, *x*_4_], reflected time spent in sleep, SB, light PA (LPA) and moderate-to-vigorous PA (MVPA), respectively. Note: when considering the raw compositional coefficients, time-use estimates and interaction effects for the complete vector of coordinates, the choice of SBP is arbitrary.

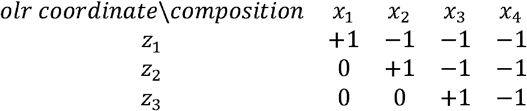

According to the sign matrix above the resultant olr-coordinates **z** = [*z*_1_, *z*_2_, *z*_3_] were then calculated as follows (ignoring the person *I* at timepoints *j* notation for simplicity)

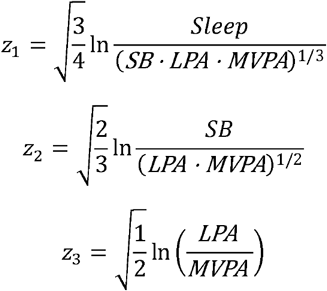

For person *i* = 1,2, …,241 and time-point *j* = 1,2, …,5, the corresponding three response (*r* = 1,2,3) *olr-coordinates z*_*rij*_were then modelled as

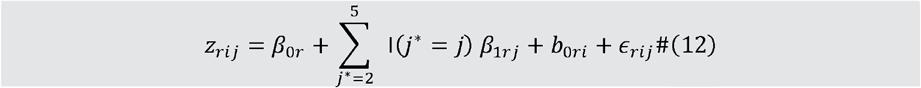

where *β*;_0*r*_is the fixed intercept specific to response olr-coordinate*r*; I(*a*) is the indicator function that is equal to 1 when the argument *a* is true, 0 otherwise; *β*_1*rj*_is the fixed ‘slope’/contrast between time-point*j* (= 2,3,4,5) and time-point 1 specific to response olr-coordinate 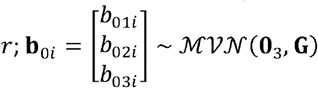 are potentially differently varying and correlated random intercepts for each response olr-coordinate *r*, specific to person *i*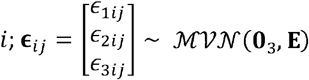are potentially differently varying and correlated random errors for each respose orl-coordinate *r*specific to person at time-point *j*; with group- and residual-side variance and covariance matrices respectively defined according Equations (9) and (10).

It is noteworthy that In the LoH study, a linear relationship between the follow-up timepoints and each olr-response cannot be assumed. However in a more general case of *J* repeated measures (or even *J*_*i*_ repeated measures per person *i*) over time that may not be equally spaced but a linear relationship between the follow-up timepoints and each olr-response, one would replace the somewhat notationally clumsy 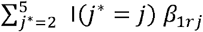 terms simply with *β*_1*r*_*t*_*ij*_for each response *r* where *t*_*ij*_is potentially continuous and unique time value for the *j*th repeated measure for person *i*. An additional level of nesting to account for nesting of participants within schools was also tested, however the variance components for this level of the model were very low, suggesting little school-to-school variation. The school-level random effects were subsequently dropped in the interest of model parsimony. In order to fit Equation (12), the three response variables were then stacked into a single column to be used as the dependent variable as described earlier. Models were fitted using R package the `nlme::lme()`^36^. R code along with alternate model specification using the `lme4::lmer()`package ^37^ are provided in supplementary material 1.

Table 2. presents parameter estimates and standard errors for the fixed effects of Equation (12). p values are reported using the ‘inner outer’ approximation of degrees of freedom as is default in `nlme::lme()` ^36^. The accurate estimation of degrees of freedom and p values in multilevel models is often discussed ^45^, however is out of scope for this article. Results suggest higher values for all three olr-coordinates over time, in particular for timepoint 3. zBMI was positively associated with *z*_3_representing the balance of LPA to MVPA suggesting the ratio of LPA to MVPA increased as zBMI increased.

**Table 2.** Fixed effect estimates for the CMRLMM

|  | **z<sub>1</sub>** |  |  |  | **z<sub>1</sub>** |  |  |  | **z<sub>1</sub>** |  |  |
| --- | --- | --- | --- | --- | --- | --- | --- | --- | --- | --- | --- |
|  | **Estimate**<br>**(S.E)** | **t-**<br>**statistic** | **p value** |  | **Estimate**<br>**(S.E)** | **t-**<br>**statistic** | **p value** |  | **Estimate**<br>**(S.E)** | **t-**<br>**statistic** | **p value** |
| **Intercept** | 0.83 (0.02) | 48.0 | <0.01 |  | 0.94 (0.04) | 22.8 | <0.01 |  | 0.93 (0.04) | 26.2 | <0.01 |
| **Timepoint**<br>**(vs. T1)** |  |  |  |  |  |  |  |  |  |  |  |
| **T2** | 0.01 (0.01) | 0.7 | 0.48 |  | 0.01 (0.02) | 0.49 | 0.63 |  | -0.01<br>(0.02) | -0.34 | 0.74 |
| **T3** | 0.09 (0.01) | 6.7 | <0.01 |  | 0.22 (0.03) | 8.5 | <0.01 |  | 0.21 (0.03) | 7.5 | <0.01 |
| **T4** | 0.03 (0.01) | 2.1 | 0.03 |  | 0.09 (0.02) | 3.8 | <0.01 |  | 0.01 (0.03) | 0.4 | 0.71 |
| **T5** | 0.04 (0.01) | 2.9 | <0.01 |  | 0.12 (0.03) | 4.6 | <0.01 |  | 0.05 (0.03) | 1.7 | 0.09 |
| **zBMI** | 0.01 (0.01) | 0.7 | 0.45 |  | 0.003<br>(0.02) | 0.2 | 0.87 |  | 0.05 (0.01) | 3.1 | <0.01 |
| **Income (vs.**<br>**middle)** |  |  |  |  |  |  |  |  |  |  |  |
| **High** | -0.001<br>(0.02) | -0.1 | 0.96 |  | 0.01 (0.05) | 0.1 | 0.92 |  | -0.07<br>(0.04) | -1.6 | 0.12 |
| **low** | 0.03 (0.02) | 1.4 | 0.16 |  | 0.10 (0.05) | 2.1 | 0.04 |  | 0.03 (0.04) | 0.8 | 0.41 |

Abbreviations: S.E = standard error; zBMI = body mass index z-score (a measure of adiposity)

The estimated level 2 random effects (between individual) variance/covariance matrix as described in Equation (9) are shown below:

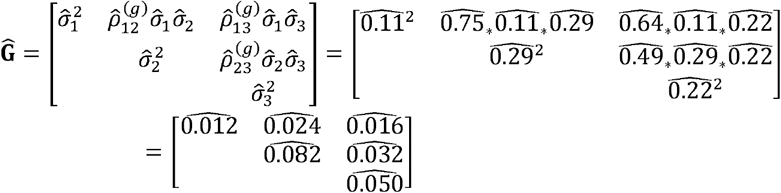

And the estimated level 1 residual (within individual and time-point) error variance/covariance matrix as described in Equation (10):

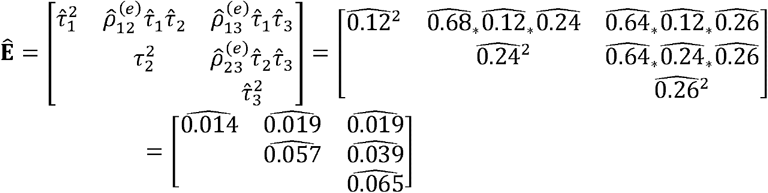

Positive correlations for all three olr-coordinates were observed in both the within- and between-person covariance matrices. Importantly, these correlations are overlooked when using the multiple univariate response modelling approach outlined earlier. Practically, this impacts our results in two ways. Firstly, the model fit statistics and statistical power will improve when compared to a multiple models approach used previously. To demonstrate this, we can fit the equivalently specified multivariate response model with the random effect and residual covariance matrices assumed to be diagonal matrices (covariances constrained to zero), referred to as the multivariate unrelated outcomes model. This model will provide the same estimates at all levels of the model as those provided by the multiple univariate models. However, because the multivariate unrelated outcomes model is nested within the fully multivariate model outlined above, it can be compared via a likelihood ratio test. Results of the likelihood ratio test suggest that the addition of the covariances between olr coordinates at both levels of the model improves model fit (Table 3).

**Table 3.** likelihood ratio test comparing CMRLMM to the multivariate unrelated outcomes model with assumed diagonal covariance matrices.

| Model | df | AIC | BIC | loglik | L ratio | p value |
| --- | --- | --- | --- | --- | --- | --- |
| **CMRLMM** | 36 | -1261 | -1048 | 666 | (1 vs 2) |  |
| **Unrelated model** | 30 | -104 | 74 | 82 | 1169 | <0.0001 |

Abbreviations: CMRLMM = compositional multivariate response linear mixed model; df = degrees of freedom; AIC = Akaike information criterion; BIC = Bayesian information criterion; loglik = log-likelihood

A second important difference is that the results for the CMRLMM will be invariant to the basis used to construct the olr-coordinates. This is not the case when constraining the covariances to be diagonal matrices as is the case when fitting independent models on each olr coordinate. To demonstrate this, we can see in Figure 1 the fitted values for 10 randomly sampled level 1 units that have been transformed back to their compositional representation when using the CMRLMM and those from multiple, independent models with olr coordinates that were constructed using two different orthonormal bases. It can be seen that the compositional representation is equivalent for the two CMRLMMs. However, results using the independent models approach differ depending on the basis used to construct the olr coordinates. Importantly, both also differ from those provided from the CMRLMM. Further model comparisons are provided in supplementary file.

**Figure 1.**
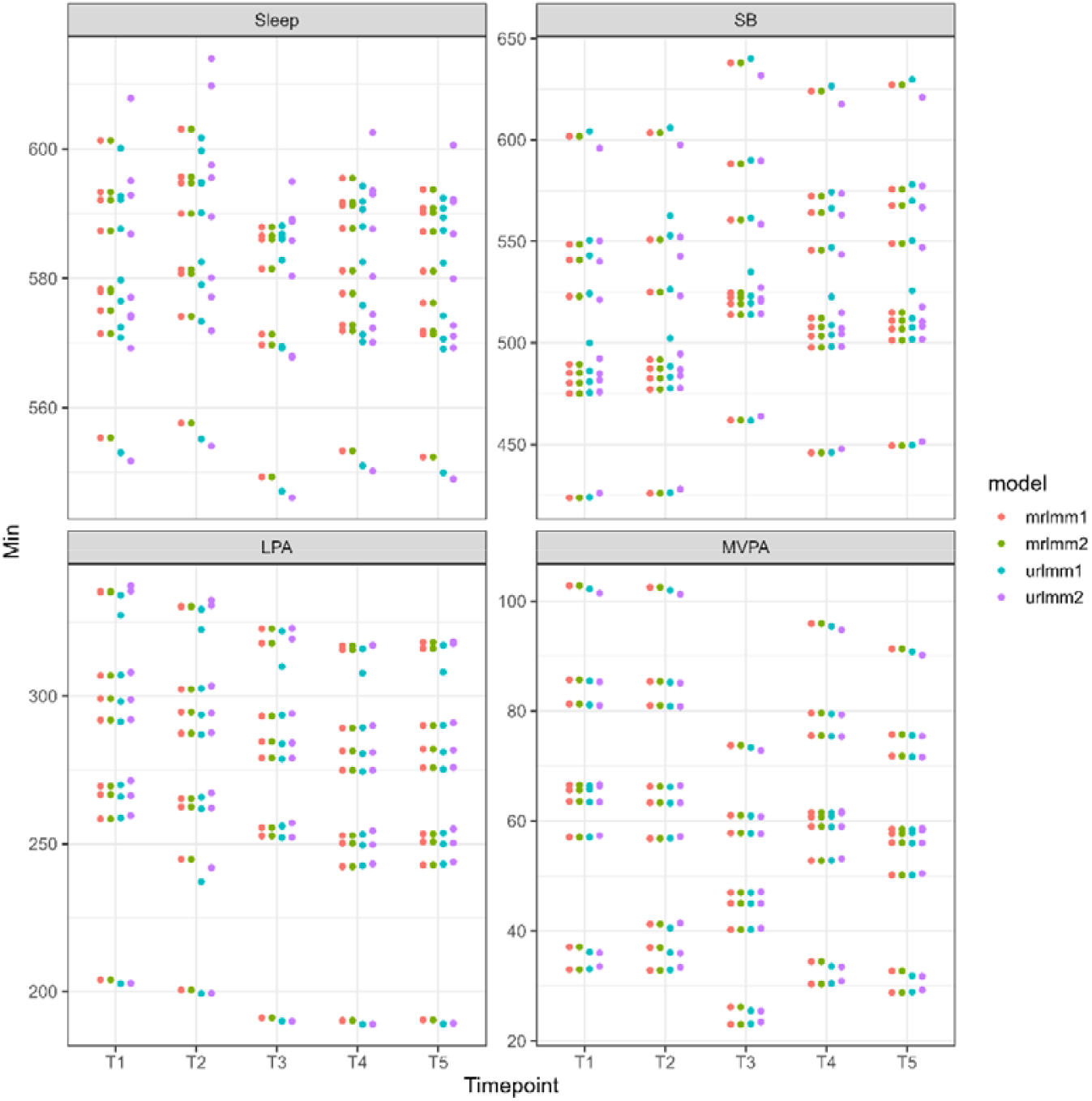
Fitted values for 10 randomly selected level 1 units with coordinates constructed using two different olr bases. Abbreviations: mrlmm = multivariate response linear mixed model; urlmm = univariate response linear mixed model

The t-statistic for each olr-coefficient are displayed in Table 2. Unlike the multiple models approach used previously, the multivariate mixed model also allows for a single test of the fixed effects for the joint effect on the vector of olr-coordinates which will be invariant to the basis chosen when constructing the olr-coordinates. The results of the multivariate F-test are presented in Table 4. Using ‘inner-outer’ approximation of denominator degrees of freedom ^46^, the F statistic for interaction between the vector of olr-coordinates and timepoints (F(12, 2475) = 9.832, p<0.001) shows that the fixed effects of the model suggest movement-behaviour composition is significantly different across timepoints. Results also suggest that there was a significant interaction between zBMI and movement-behaviour composition (F(3,2475) = 4.44, p=0.004), whereas parental income and movement-behaviour composition were not significantly associated (F(6,2475) = 1.86, p=0.08). The ability to perform multivariate tests such as these is limited when creating separate models for individual olr coordinates. Three-way interactions between olr-coordinates, timepoint and covariates were also tested, but were not significant, suggesting children followed a similar pattern of across all timepoints.

**Table 4.**
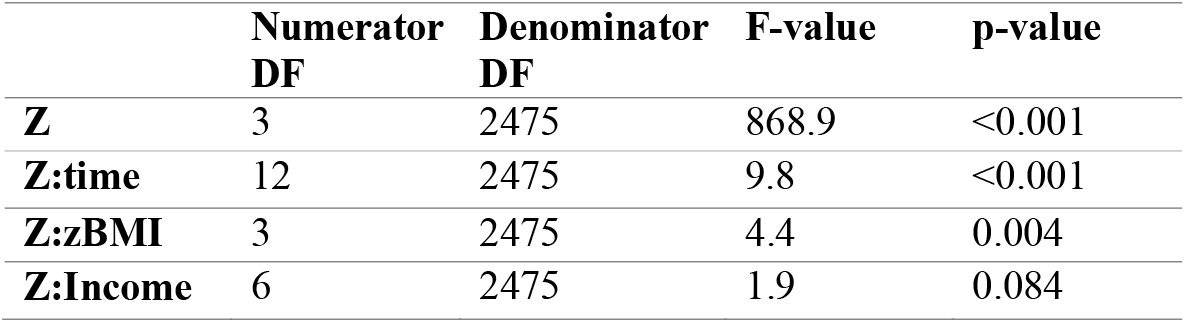
Multivariate F test for the CMRLMM

While the results of the multivariate F-test suggest movement-behaviour compositions are different across timepoints, they do not give an indication of which timepoints differ, on which components in what direction. In order to investigate this, the estimates presented in Table 2 which are specific to the basis chosen when constructing the olr-coordinates **z** = [*z*_1_,*z*_2_,*z*_3_], need to be back-transformed to their corresponding raw compositional representation **x** = [*x*_1_, *x*_2_, *x*_3,_*x*_4_], using **β**_*x*_= olr ^−1^ (β_*z*_). For example, back transforming the vector of intercept estimates 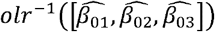, will result in the estimate mean movement-behaviour composition at baseline (assuming zBMI of zero and reference SES group), as below

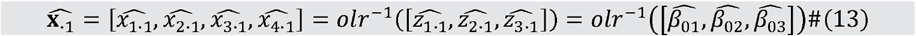

Once transformed into the simplex space, the compositional coefficient estimates are now invariant to the basis chosen when constructing the olr-coordinates for both the fixed effects and random effects. After back transforming the estimates for timepoints T2 to T5, the compositional coefficients can be interpreted as the perturbation vector for that timepoint when applied to the baseline composition (which has been closed to 1440 min/day in Table 5) to obtain estimates for a given timepoint. The perturbation vector can be interpreted substantively as relative reallocations for each behaviour when compared to the neutral perturbation vector, subject to closure as outlined in Section 1.1. For example, with a four-part composition, as in our demonstration, the neutral perturbation vector [1/D, …, 1/D] is [0.25, 0.25, 0.25, 0.25]. When the compositional coefficients for timepoints 2-5 are compared to the neutral perturbation vector we can see that in our example dataset time appears to be reallocated away from MVPA (values <0.25) and towards SB (values >0.25) as children age across the timepoints, with the largest changes occurring during T3 (the school holiday period).

**Table 5.** Compositional coefficient estimates for fixed effects.

| Timepoint | Sleep | SB | LPA | MVPA |
| --- | --- | --- | --- | --- |
| **Intercept**<br>$\hat{\beta}_0$ | 584 | 483 | 294 | 79 |
| **T2** | 0.252 | 0.251 | 0.247 | 0.250 |
| **T3** | 0.266 | 0.289 | 0.256 | 0.190 |
| **T4** | 0.255 | 0.267 | 0.240 | 0.237 |
| **T5** | 0.257 | 0.271 | 0.243 | 0.228 |

While compositional coefficients are meaningful, it may be preferred to make model-based point estimates for the compositions, and differences, across timepoints using the fixed effects from the CMRLMM. Figure 2 provides estimated movement-behaviour compositions at the five timepoints (for a participant of mean zBMI and income category) via predicted olr-values that have been back transformed into the simplex space *S*^4^, 95% percentile intervals for the compositions were estimated using non-parametric ‘cases’ bootstrapping procedure with 1000 replicates, which resamples level-2 observations (participants) ^47,48^.

**Figure 2.**
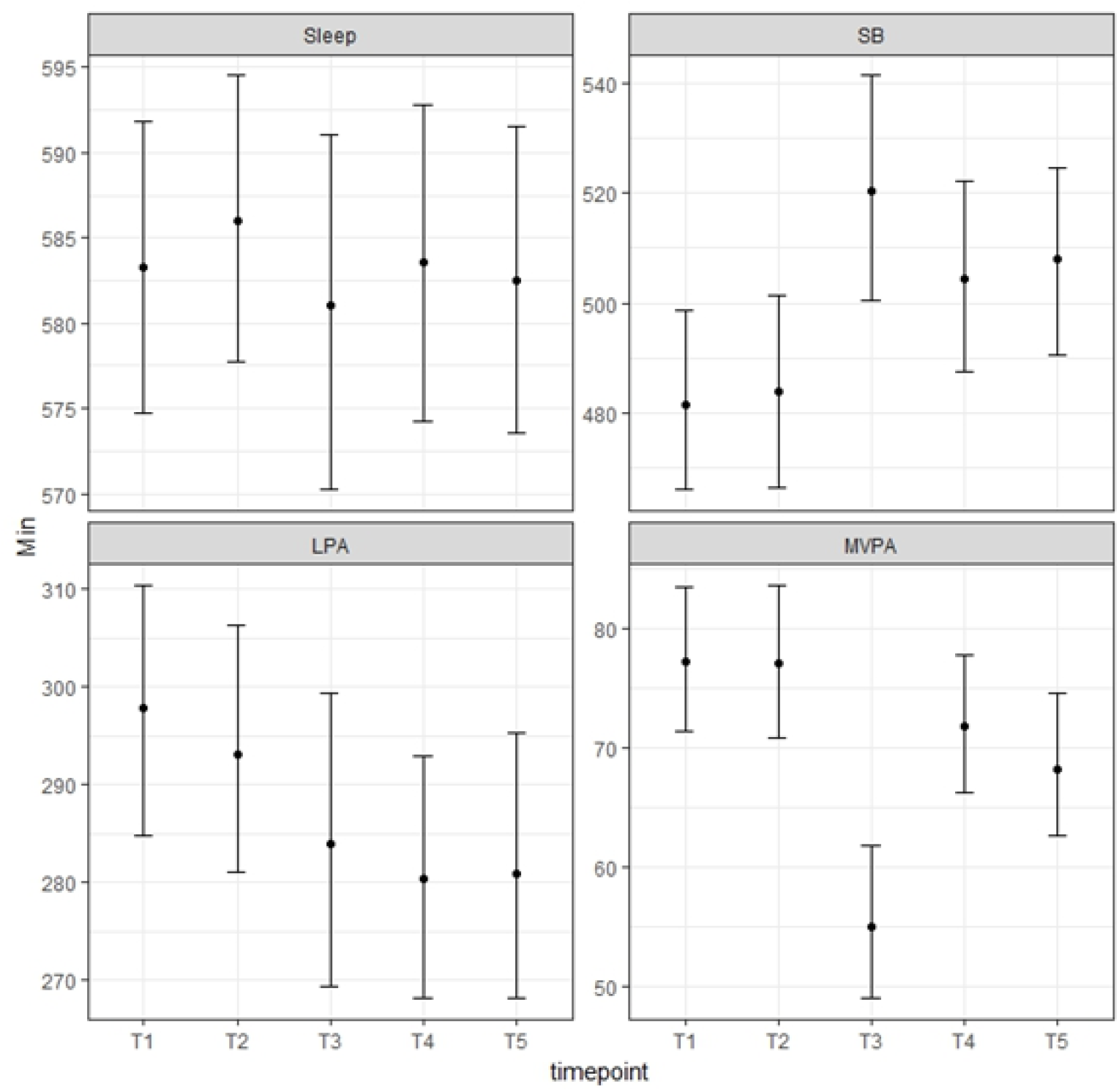
Estimated movement-behaviour compositions for an average participant across the five timepoints. Sleep: range 581-586 min/day. SB: range 481-520 min/day. LPA: range 280-298 min/day. MVPA: range 55-77 min/day.

We can see that Timepoint 3 during the school-holiday period appears to have the largest differences, particularly in relation to the time spent in SB and MVPA. It is also noteworthy that despite timepoints 3,4 and 5 being significantly and positively associated with (Table 2), suggesting more sleep across these timepoints, we can see that the proportion of sleep is relatively stable across all timepoints, and in fact estimated to be lower in timepoints T3 and T5 than at baseline. The seemingly contradictory results are due to the way must be interpreted. As mentioned in Section 1.1, the first pivot coordinate represents the dominance of sleep relative to the geometric mean of SB, LPA and MVPA. Despite sleep remaining relatively stable across all timepoints, there is a significant change in across timepoints due to the change in the sub-composition of the remaining behaviours (increased contribution of SB and decreased MVPA). This demonstrates why the CMRLMM is preferred to the previously used method of trying to draw inferences about time spent in each behaviour from pivot coordinates in univariate models as has been done previously ^29,31^. In order to estimate differences between timepoints we can use the approach suggested by Martín Fernández, Daunis-i-Estadella ^49^ to determine group differences. Here, we use the fixed effects of the model to calculate the log-ratio difference in the predicted movement-behaviour compositions between timepoint 3 and other timepoints with bootstrapped percentile intervals as shown in Figure 3.

**Figure 3.**
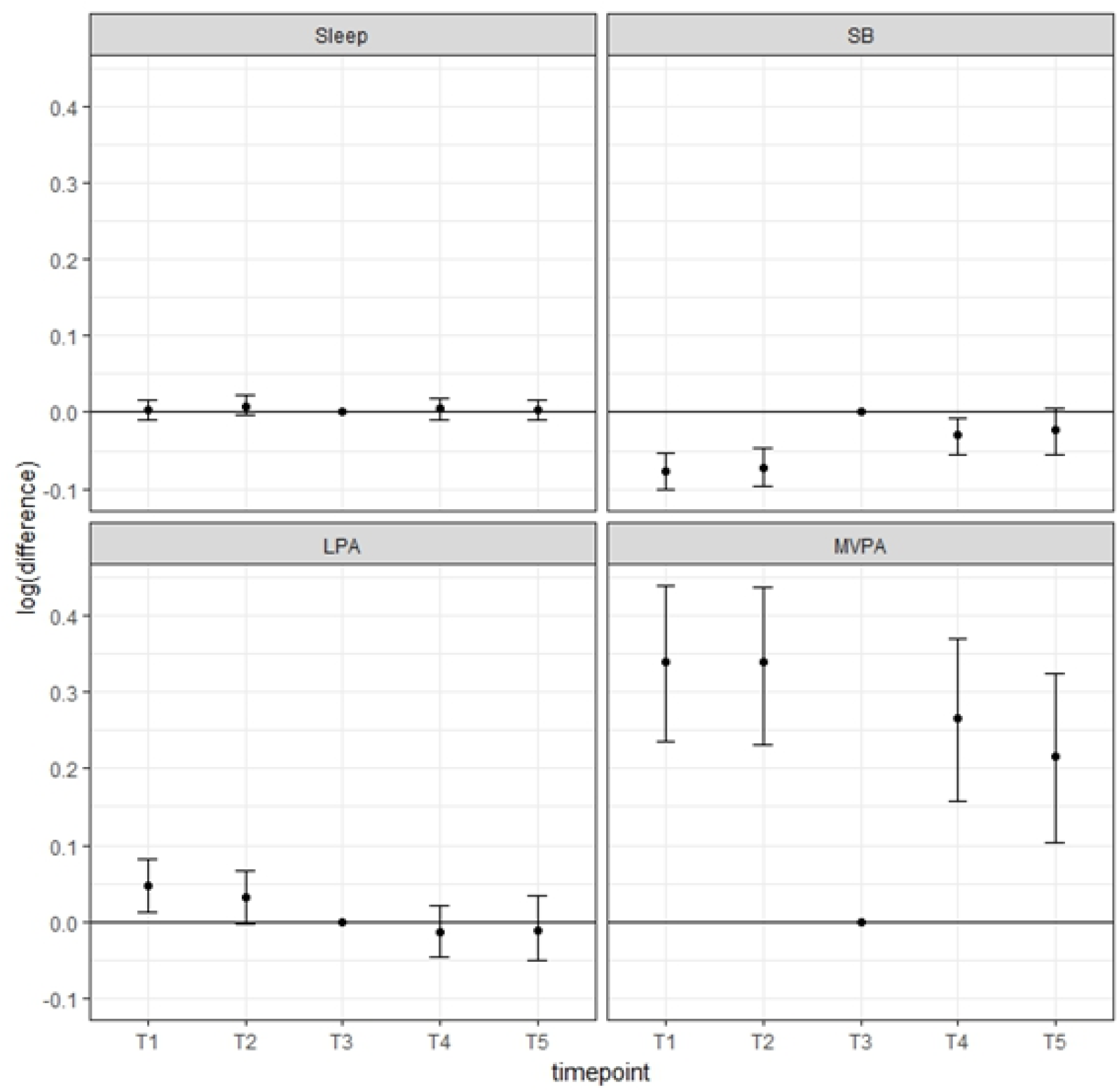
Estimated log-ratio difference in compositional parts for each timepoint when compared to timepoint 3 (school holidays). Note: values above the horizontal line indicate relatively higher proportions of that component during the in-school timepoint compared to the school-holiday period; values below the horizontal line indicate relatively lower proportions of that component during the in-school timepoint compared to the school-holiday period

These findings suggest that children’s movement behaviour compositions change as they age, with distinct differences evident during the school-holiday period. These changes are characterised by lower contributions of MVPA and higher contributions of SB. Given the unfavourable associations reported between reallocating time away from MVPA to SB ^1^, this indicates the school holiday period may be a key intervention point for future public health initiatives.

## 4. Discussion

Key strengths of the CMRLMM outlined in this paper is its ability to include all daily time-use components as dependent variables in a single analytical model, within a multi-level framework. A multi-level framework is relevant for many epidemiological applications such as the analysis of repeated measures of movement behaviours over time (e.g., time-use intervention studies or longitudinal cohort studies of time use) or the analysis of time-use behaviours obtained by a clustered sampling process (e.g., students sampled from within classes, which are nested within schools, or patients sampled from within hospitals). A key benefit of the CMRLMM is that it provides consistent results regardless of how the compositional time-use behaviours are ordered in the model. The CoDA log-ratio transformation overcomes the issue of perfect multi-collinearity between the time-use components, and the variable stacking procedure outlined in this paper enables all the log ratios to be considered as dependent variables simultaneously. Another strength of the analytical pipeline presented in this paper is the interpretability of the model log-ratio coefficients in the original compositional units (minutes/day) via a back-transformation. As we have demonstrated, specific research questions regarding meaningful differences in time use (e.g., comparing children’s time use during school terms to the holidays) can be explored by post-hoc comparisons, and hypotheses via bootstrapping.

There are limitations to the CMRLMM which should be considered. First, the CoDA olr transformation cannot be implemented if there are zero values in any of the time-use components, as the logarithm of zero is undefined. However, this is true of all CoDA techniques, and there are published methods for dealing with zero values in compositional variables, which are beyond the scope of this paper. In terms of the implementation of the CMRLMM in the R environment, it should be considered that these models can have long run times, and convergence issues can arise. However, these issues can be addressed by following recommended troubleshooting procedures (e.g., rescaling and centring continuous variables, trying various optimisers, increasing the maximum number of iterations) ^45^. Importantly, the CRLMM expressed in compositional coefficients is not more complicated than univariate response models used previously, so the benefits justify the greater computational complexity.

The use of CoDA in time-use epidemiology has grown at a rapid rate in recent years, leading to a paradigm shift in the way that people view the relationship between time use and health. For example, the acceptance that all time-use behaviours are interrelated and jointly contribute to health has led to various countries adopting integrated 24-hour movement behaviour guidelines that promote an optimal mix of these behaviours for different age groups ^50-52^. Similarly, multi-component intervention studies now accept the co-dependent nature of time use and aim to simultaneously change time spent in multiple behaviours ^7^. Our proposed CMRLMM provides the tools required to allow researchers to better understand how time use changes longitudinally due to natural interventions (such as in the example given in this paper), or in experimental designs. The CMRLMM approach will also support analyses that explore how different personal and sociodemographic factors may be related to different time reallocation patterns. This knowledge will allow more targeted public health initiatives that may aim to block unfavourable reallocations (such as from MVPA to SB as in the current example), or alternatively nudging people to make favourable reallocations.

## Supporting information

MRLMM for CoDa example

MRLMM for CoDa model comparison

Mathematical background

## Data Availability

All data produced in the present study are available upon reasonable request to the authors

## 5. Statements and declarations

## Ethical considerations

Ethical approval was obtained from The University of South Australia Human Research Ethics Committee (200980), the South Australian Department of Education and Child Development (2008–0055) and the Adelaide Catholic Education Centre (201820) for the original Life on Holidays study.

## References

1. Miatke A, Olds T, Maher C, et al. The association between reallocations of time and health using compositional data analysis: a systematic scoping review with an interactive data exploration interface. International Journal of Behavioral Nutrition and Physical Activity. 2023;20(1):127.

2. Warburton DE, Bredin SS. Health benefits of physical activity: a systematic review of current systematic reviews. Current opinion in cardiology. 2017;32(5):541–556.

3. Müller-Riemenschneider F, Reinhold T, Nocon M, Willich SN. Long-term effectiveness of interventions promoting physical activity: a systematic review. Preventive medicine. 2008;47(4):354–368.

4. Prince S, Saunders T, Gresty K, Reid R. A comparison of the effectiveness of physical activity and sedentary behaviour interventions in reducing sedentary time in adults: a systematic review and metaOanalysis of controlled trials. Obesity reviews. 2014;15(11):905–919.

5. Saunders TJ, McIsaac T, Douillette K, et al. Sedentary behaviour and health in adults: an overview of systematic reviews. Applied Physiology, Nutrition, and Metabolism. 2020;45(10):S197–S217.

6. Albakri U, Drotos E, Meertens R. Sleep health promotion interventions and their effectiveness: an umbrella review. International Journal of Environmental Research and Public Health. 2021;18(11):5533.

7. Lewis BA, Napolitano MA, Buman MP, Williams DM, Nigg CR. Future directions in physical activity intervention research: expanding our focus to sedentary behaviors, technology, and dissemination. Journal of behavioral medicine. 2017;40:112–126.

8. Pedišić Ž. Measurement issues and poor adjustments for physical activity and sleep undermine sedentary behaviour research—the focus should shift to the balance between sleep, sedentary behaviour, standing and activity. Kinesiology. 2014;46(1.):135–146.

9. Dumuid D, Stanford TE, Martin-Fernández J-A, et al. Compositional data analysis for physical activity, sedentary time and sleep research. Statistical methods in medical research. 2018;27(12):3726–3738.

10. von Rosen P. Analysing time-use composition as dependent variables in physical activity and sedentary behaviour research: different compositional data analysis approaches. Journal of Activity, Sedentary and Sleep Behaviors. 2023;2(1):23.

11. Aitchison J. The statistical analysis of compositional data. Journal of the Royal Statistical Society: Series B (Methodological). 1982;44(2):139–160.

12. Aitchison J. A concise guide to compositional data analysis. 2005:

13. Egozcue JJ, PawlowskyOGlahn V. Basic concepts and procedures. Compositional data analysis: Theory and applications. 2011:12–28.

14. MateuOFigueras G, PawlowskyOGlahn V, Egozcue JJ. The principle of working on coordinates. Compositional data analysis: Theory and applications. 2011:29–42.

15. Tolosana-Delgado R, Van den Boogaart K. Linear models with compositions in R. Compositional data analysis: theory and applications. 2011;

16. Egozcue JJ, Pawlowsky-Glahn V. Compositional data: the sample space and its structure. Test. 2019;28(3):599–638.

17. Martín-Fernández J. Comments on: Compositional data: the sample space and its structure. Test. 2019;28(3):653–657.

18. Egozcue JJ, Pawlowsky-Glahn V, Mateu-Figueras G, Barcelo-Vidal C. Isometric logratio transformations for compositional data analysis. Mathematical geology. 2003;35(3):279–300.

19. Martín-Fernández JA, Pawlowsky-Glahn V, Egozcue JJ, Tolosona-Delgado R. Advances in principal balances for compositional data. Mathematical Geosciences. 2018;50:273–298.

20. Martín-Fernández JA, Donato VD, Pawlowsky-Glahn V, Egozcue JJ. Insights in Hierarchical Clustering of Variables for Compositional Data. Mathematical Geosciences. 2024;56(3):415–435.

21. Egozcue JJ, Pawlowsky-Glahn V. Groups of parts and their balances in compositional data analysis. Mathematical Geology. 2005;37:795–828.

22. McGregor DE, Dall PM, Palarea-Albaladejo J, Chastin SF. Compositional Data Analysis in Physical Activity and Health Research. Looking for the Right Balance. Advances in Compositional Data Analysis: Festschrift in Honour of Vera Pawlowsky-Glahn. Springer; 2021:363–382.

23. Fišerová E, Hron K. On the interpretation of orthonormal coordinates for compositional data. Mathematical Geosciences. 2011;43:455–468.

24. Van den Boogaart KG, Tolosana-Delgado R. Analyzing compositional data with R. vol 122. Springer; 2013.

25. Dumuid D, Pedišić Ž, Stanford TE, et al. The compositional isotemporal substitution model: a method for estimating changes in a health outcome for reallocation of time between sleep, physical activity and sedentary behaviour. Statistical methods in medical research. 2019;28(3):846–857.

26. Ben Bolker JP, Emi Tanaka, Phillip Alday, Wolfgang Viechtbauer. CRAN Task View: Mixed, Multilevel, and Hierarchical Models in R. Accessed 20th May, 2024. https://cran.r-project.org/web/views/MixedModels.html

27. Schabenberger O. Introducing the GLIMMIX procedure for generalized linear mixed models. SUGI 30 Proceedings. 2005;196:1–20.

28. Achana F, Gallacher D, Oppong R, et al. Multivariate generalized linear mixed-effects models for the analysis of clinical trial–based cost-effectiveness data. Medical Decision Making. 2021;41(6):667–684.

29. Larisch L-M, Bojsen-Møller E, Nooijen CF, et al. Effects of two randomized and controlled multi-component interventions focusing on 24-hour movement behavior among office workers: a compositional data analysis. International Journal of Environmental Research and Public Health. 2021;18(8):4191.

30. Larsson K, Von Rosen P, Rossen J, Johansson U-B, Hagströmer M. Relative time in physical activity and sedentary behaviour across a 2-year pedometer-based intervention in people with prediabetes or type 2 diabetes: a secondary analysis of a randomised controlled trial. Journal of Activity, Sedentary and Sleep Behaviors. 2023;2(1):10.

31. Pasanen J, Leskinen T, Suorsa K, et al. Effects of physical activity intervention on 24-h movement behaviors: a compositional data analysis. Scientific Reports. 2022;12(1):8712.

32. Suorsa K, Leskinen T, Pasanen J, et al. Changes in the 24-h movement behaviors during the transition to retirement: compositional data analysis. International Journal of Behavioral Nutrition and Physical Activity. 2022;19(1):121.

33. Farcomeni A. A review of modern multiple hypothesis testing, with particular attention to the false discovery proportion. Statistical methods in medical research. 2008;17(4):347–388.

34. Greenacre M. Towards a pragmatic approach to compositional data analysis. 2017;

35. Hox J, Moerbeek M, Van de Schoot R. Multilevel analysis: Techniques and applications. Routledge; 2017.

36. Pinheiro JC, Bates DM. Linear mixed-effects models: basic concepts and examples. Mixed-effects models in S and S-Plus. 2000:3–56.

37. Bates D, Maechler M, Bolker B, et al. Package ‘lme4’. convergence. 2015;12(1):2.

38. Curran PJ, McGinley JS, Serrano D, Burfeind C. A multivariate growth curve model for three-level data. 2012;

39. Twisk JW. Applied multilevel analysis: a practical guide for medical researchers. Cambridge university press; 2006.

40. Baldwin SA, Imel ZE, Braithwaite SR, Atkins DC. Analyzing multiple outcomes in clinical research using multivariate multilevel models. Journal of consulting and clinical psychology. 2014;82(5):920.

41. Snijders TA, Bosker R. Multilevel analysis: An introduction to basic and advanced multilevel modeling. 2011;

42. Watson A, Maher C, Tomkinson GR, et al. Life on holidays: study protocol for a 3-year longitudinal study tracking changes in children’s fitness and fatness during the in-school versus summer holiday period. BMC Public Health. 2019;19:1–8.

43. Phillips LR, Parfitt G, Rowlands AV. Calibration of the GENEA accelerometer for assessment of physical activity intensity in children. Journal of science and medicine in sport. 2013;16(2):124–128.

44. Van Hees VT, Sabia S, Anderson KN, et al. A novel, open access method to assess sleep duration using a wrist-worn accelerometer. PloS one. 2015;10(11):e0142533.

45. Bolker B. GLMM FAQ. Accessed January 31st, 2025. bbolker.github.io/mixedmodels-misc/glmmFAQ

46. Pinheiro J, Bates D. Mixed-effects models in S and S-PLUS. Springer science & business media; 2006.

47. Carpenter JR, Goldstein H, Rasbash J. A novel bootstrap procedure for assessing the relationship between class size and achievement. Journal of the Royal Statistical Society Series C: Applied Statistics. 2003;52(4):431–443.

48. Leeden Rvd, Meijer E, Busing FM. Resampling multilevel models. Handbook of multilevel analysis. Springer; 2008:401–433.

49. Martín Fernández JA, Daunis-i-Estadella P, Mateu i Figueras G. On the interpretation of differences between groups for compositional data. SORT: statistics and operations research transactions, 2015, vol 39, núm 2, p 231–252. 2015;

50. Ross R, Chaput J-P, Giangregorio LM, et al. Canadian 24-hour movement guidelines for adults aged 18–64 years and adults aged 65 years or older: an integration of physical activity, sedentary behaviour, and sleep. Applied Physiology, Nutrition, and Metabolism. 2020;45(10):S57–S102.

51. Draper CE. The South African 24-hour movement guidelines for birth to 5 years. South African Journal of Child Health. 2021;15(2):58–59.

52. Okely AD, Ghersi D, Hesketh KD, et al. A collaborative approach to adopting/adapting guidelines-The Australian 24-Hour Movement Guidelines for the early years (Birth to 5 years): an integration of physical activity, sedentary behavior, and sleep. BMC public health. 2017;17:167–190.

