## Supplementary material for "When the outcome is compositional - a method for conducting compositional response linear mixed models for physical activity, sedentary behaviour and sleep research": MRLMM for CoDa example

```
library(compositions)
library(lme4)
library(forcats)
library(ggplot2); theme_set(theme_bw())
library(plyr)
library(tidyr)
library(dplyr)
library(nlme)
library(broom.mixed)
library(lattice)
library(gridExtra)
```

Import data and make compositions and olrs

```
#make a copy to work with
D <- data
# have a quick look at dataset
head(D)
```

```
##         id       Zbmi Income.cat sleep    sb   lpa  mvpa timepoint
## 1 LOH002C  1.5954419       High 538.1 441.0 373.0  70.6        T1
## 2 LOH003C -1.6143211       <NA> 555.1 556.4 238.6  65.6        T1
## 3 LOH004C  0.1637041        Low 568.6 456.3 264.9 139.1        T1
## 4 LOH005C  1.0277690        Low 561.9 526.7 240.0  52.0        T1
## 5 LOH006C  0.3445147     Medium 571.1 613.7 194.9  53.0        T1
## 6 LOH007C  0.1579152       <NA> 555.6 446.4 325.3  56.6        T1
```

```
# make comp and adjust to sum to 1440
D$a.comp <-clo(acomp(cbind.data.frame(D$sleep, D$sb, D$lpa, D$mvpa)), total=1440)
#add the accelerometry compositions to the dataset as separate variables
D$Sleep <- D$a.comp[,1]
D$SB <- D$a.comp[,2]
D$LPA <- D$a.comp[,3]
D$MVPA <- D$a.comp[,4]
# remove vars that are not closed for simplicity
D <-  D %>% select(-c(sleep, sb,lpa,mvpa))
# make ilrs
# create copy of data
d <-  D
#sign matrix in manuscript
sm <-  matrix(c(1, -1, -1, -1,
                0, 1, -1, -1,
                0, 0, 1, -1),
              nrow = 3,
              ncol = 4,
```

```
                byrow = TRUE)
#basis
SBP <- gsi.buildilrBase(t(sm))
d$ilrs <- ilr(d$a.comp, V = SBP)
d$ilr1 <- d$ilrs[,1] # assign each to dataframe individually
d$ilr2 <- d$ilrs[,2]
d$ilr3 <- d$ilrs[,3]
```

##### Restructure dataframe

To fit multivariate model we need to restructure so that vector of olrs are in single column along with dummy codes to be able to switch on and off

```
# pivot dataframe
long <-  d %>% mutate(obs=factor(1:n())) %>%
  pivot_longer(c(ilr1:ilr3), names_to = "ilr.no", values_to = "value")
```

Let's have a look at original and new data structure

```
#vars we might want to have a look at the various dataframes
peek <-  c("id", "Zbmi", "timepoint", "ilr1", "ilr2", "ilr3", "obs", "ilr.no", "value")
#original wide format
d %>% select(any_of(peek)) %>% head()
```

```
##        id       Zbmi timepoint      ilr1      ilr2      ilr3
## 1 LOH002C  1.5954419        T1 0.7495392 0.8162848 1.1770134
## 2 LOH003C -1.6143211        T1 0.8595571 1.2184622 0.9130253
## 3 LOH004C  0.1637041        T1 0.6904635 0.7069862 0.4554894
## 4 LOH005C  1.0277690        T1 0.9513168 1.2661329 1.0814457
## 5 LOH006C  0.3445147        T1 0.9758398 1.4681563 0.9207907
## 6 LOH007C  0.1579152        T1 0.8770452 0.9723143 1.2365451
```

```
#long format
long %>% select(any_of(peek)) %>% head()
```

```
## # A tibble: 6 x 6
##   id       Zbmi timepoint obs   ilr.no value
##   <fct>   <dbl> <fct>     <fct> <chr>  <dbl>
## 1 LOH002C  1.60 T1        1     ilr1   0.750
## 2 LOH002C  1.60 T1        1     ilr2   0.816
## 3 LOH002C  1.60 T1        1     ilr3   1.18
## 4 LOH003C -1.61 T1        2     ilr1   0.860
## 5 LOH003C -1.61 T1        2     ilr2   1.22
## 6 LOH003C -1.61 T1        2     ilr3   0.913
```

```
#note: when looking at the obs var above, it seems to just repeat the id var.
# but the obs var will be different for each timepoint, it essentially just groups the 3 ilrs together
#e.g.,
long %>% filter(id == "LOH002C") %>%  select(any_of(peek))
```

```
## # A tibble: 15 x 6
##    id       Zbmi timepoint obs   ilr.no value
##    <fct>   <dbl> <fct>     <fct> <chr>  <dbl>
##  1 LOH002C  1.60 T1        1     ilr1   0.750
##  2 LOH002C  1.60 T1        1     ilr2   0.816
##  3 LOH002C  1.60 T1        1     ilr3   1.18
##  4 LOH002C  1.60 T2        318   ilr1   0.913
##  5 LOH002C  1.60 T2        318   ilr2   1.21
##  6 LOH002C  1.60 T2        318   ilr3   1.53
##  7 LOH002C  1.60 T3        604   ilr1   0.712
##  8 LOH002C  1.60 T3        604   ilr2   0.903
##  9 LOH002C  1.60 T3        604   ilr3   1.25
## 10 LOH002C  1.60 T4        793   ilr1   0.778
## 11 LOH002C  1.60 T4        793   ilr2   0.972
## 12 LOH002C  1.60 T4        793   ilr3   1.18
## 13 LOH002C  1.60 T5        1036  ilr1   0.862
## 14 LOH002C  1.60 T5        1036  ilr2   1.24
## 15 LOH002C  1.60 T5        1036  ilr3   1.30
```

By treating the new dummy variable as a factor we functionally achieve the same results as if specifying D-1 individual dummy variables as demonstrated in the manuscript, yet the data structure and code are simpler to interpret.

### Multivariate mixed model

Brief description of each part of model call

##### Fixed effects

For the fixed effects we will include our dummy var column indicating which olr response (ilr.no).

Then the interaction between this column and other variables of interest. To start with we'll just look at changes over time with no covariates

We'll also remove the overall intercept (-1 in model call), because we want a separate intercept for each olr response, not differences between olrs

##### Random effects

For the random effects we'll use similar coding (ilr.no - 1) to allow (correlated) random intercept for each olr response

##### Residual error

We also need to include a few other things to allow multivariate residual structure specified in manuscript

varIdent allows for heterogeneous residual variance for each olr

corSymm allows for correlations among olr terms. Note obs indicator variable nested within id for this call. This is equivalent to nesting the timepoint, which simply groups each complete vector of olr coordinates and assumes a homogeneous variance across timepoints

```r
# specifiying control arguments for nlme::lme here to use in all subsequent mods
control <-  lmeControl(opt='optim', maxIter = 200, msMaxIter = 200, msMaxEval = 200)
# we're increasing iterations to ensure convergence
# run model
(mod.lme <-  nlme::lme(value ~ -1 + ilr.no + ilr.no:timepoint,
                        random = ~ ilr.no - 1 | id,
                        weights = varIdent(form = ~ 1 | ilr.no),
                        correlation = corSymm(form = ~1 | id/obs),
                        control = control,
                        data=long))
```

```
## Linear mixed-effects model fit by REML
##   Data: long
##   Log-restricted-likelihood: 661.023
##   Fixed: value ~ -1 + ilr.no + ilr.no:timepoint
##             ilr.noilr1             ilr.noilr2             ilr.noilr3
##            0.863602312            1.020861954            0.984494752
## ilr.noilr1:timepointT2 ilr.noilr2:timepointT2 ilr.noilr3:timepointT2
##            0.004643064            0.017258910           -0.013352770
## ilr.noilr1:timepointT3 ilr.noilr2:timepointT3 ilr.noilr3:timepointT3
##            0.091650045            0.239307905            0.236739644
## ilr.noilr1:timepointT4 ilr.noilr2:timepointT4 ilr.noilr3:timepointT4
##            0.023392000            0.082746221            0.005346990
## ilr.noilr1:timepointT5 ilr.noilr2:timepointT5 ilr.noilr3:timepointT5
##            0.024586447            0.107881370            0.048400999
##
## Random effects:
##  Formula: ~ilr.no - 1 | id
##  Structure: General positive-definite, Log-Cholesky parametrization
##            StdDev    Corr
## ilr.noilr1 0.1217304 ilr.n1 ilr.n2
## ilr.noilr2 0.2982309 0.762
## ilr.noilr3 0.2525221 0.695  0.583
## Residual   0.1311488
##
## Correlation Structure: General
##  Formula: ~1 | id/obs
##  Parameter estimate(s):
##  Correlation:
##   1     2
## 2 0.716
## 3 0.646 0.634
## Variance function:
##  Structure: Different standard deviations per stratum
##  Formula: ~1 | ilr.no
##  Parameter estimates:
##     ilr1     ilr2     ilr3
## 1.000000 1.918127 1.978729
## Number of Observations: 3711
## Number of Groups: 349
```

Model specs look ok

```r
#model diagnostics
diagnostics <- data.frame(r = resid(mod.lme, type = "normalized"), f = fitted(mod.lme),
                          ilr.no = long$ilr.no)
#
plot(x=diagnostics$f,y=diagnostics$r, col = factor(diagnostics$ilr.no))
```

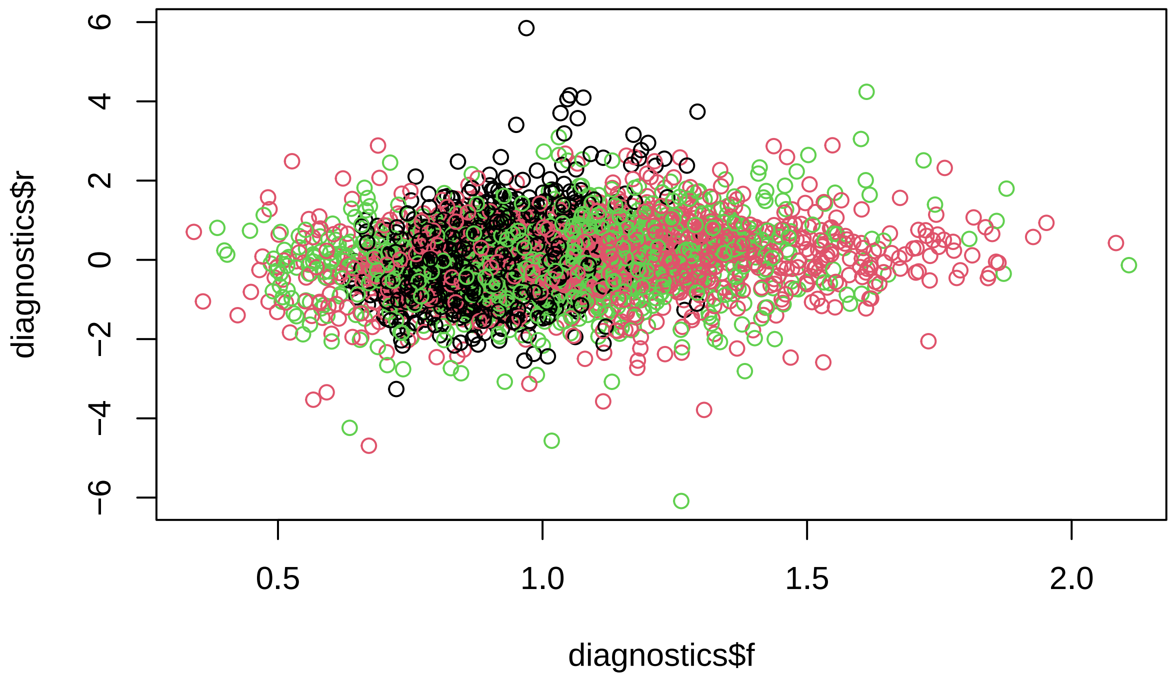


```r
## overal QQ plot
qqmath(~r, data = diagnostics, distribution = qnorm)
```

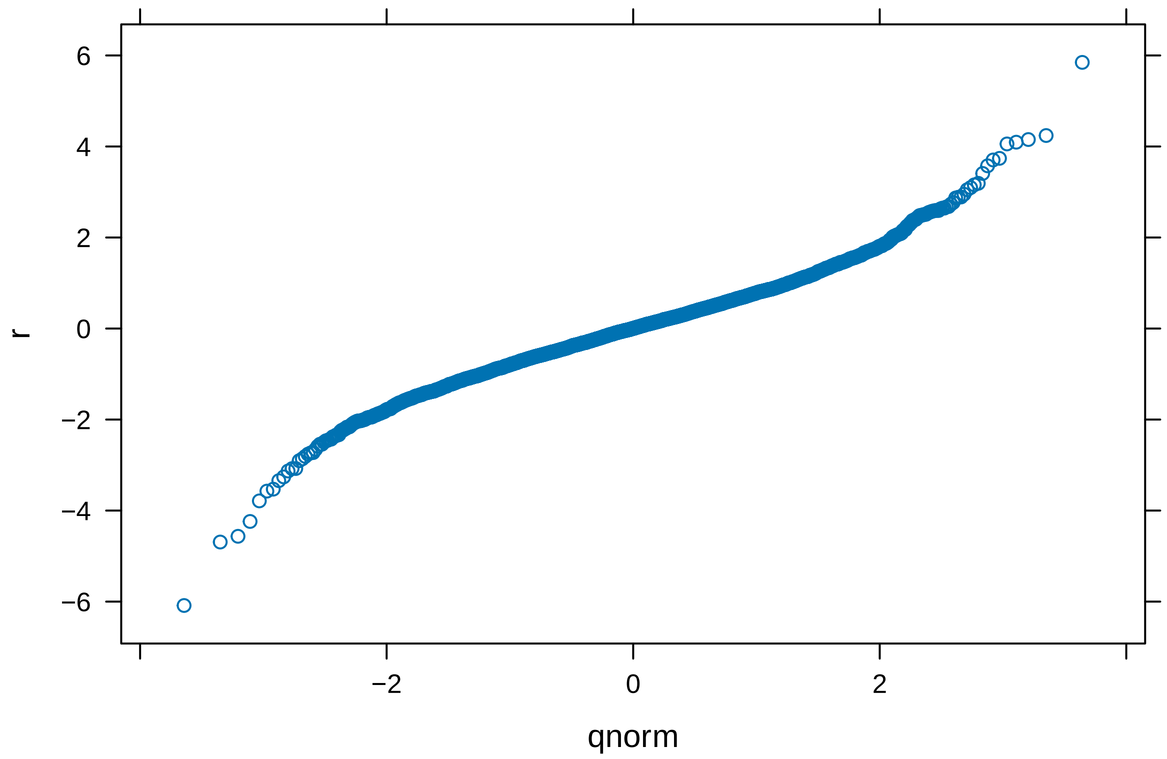


```
# and split by ilr no
qqmath(~r | ilr.no, data = diagnostics, distribution = qnorm, prepanel = prepanel.qqmathline,
       panel = function(x, ...) {
         panel.qqmathline(x, ...)
         panel.qqmath(x, ...)
       })
```

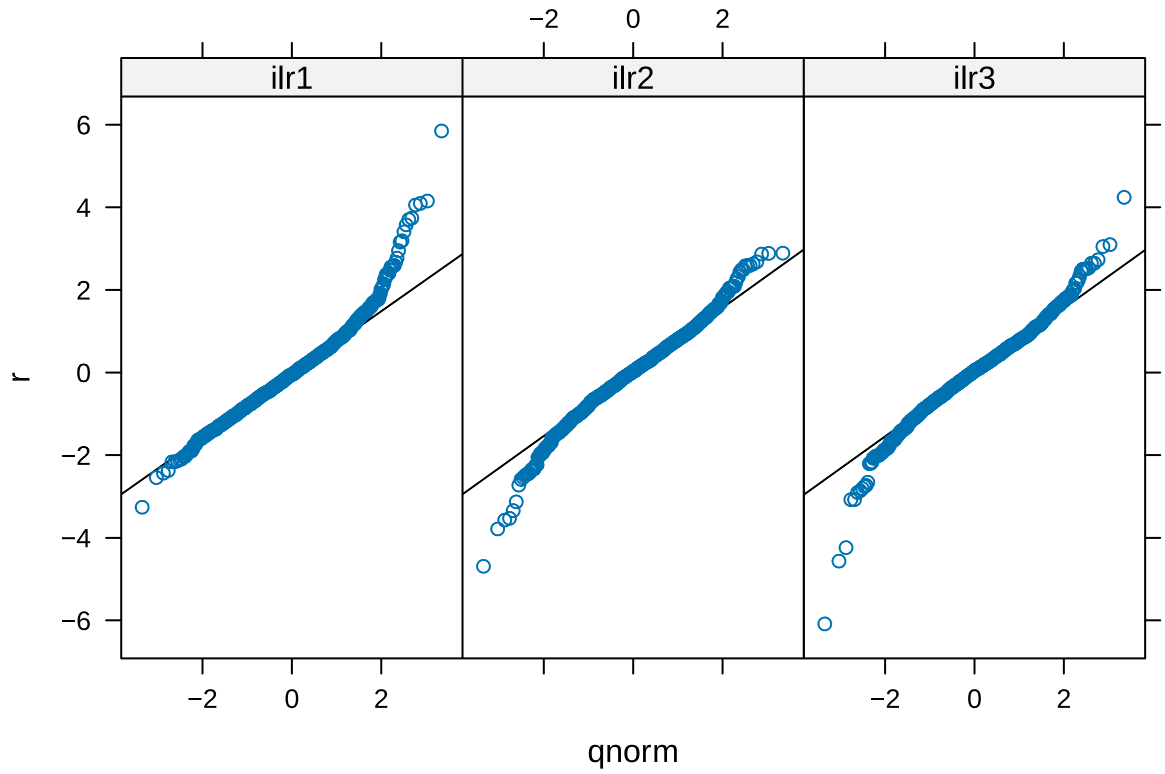


```
#and random effects
ran.diag <- ranef(mod.lme)
colnames(ran.diag) <- c("ilr1.int", "ilr2.int", "ilr3.int")
#plots for random intercepts
qq1 <- qqmath(~ilr1.int, data = ran.diag, distribution = qnorm)
qq2 <- qqmath(~ilr2.int, data = ran.diag, distribution = qnorm)
qq3 <- qqmath(~ilr3.int, data = ran.diag, distribution = qnorm)
grid.arrange(qq1,qq2,qq3, ncol=3)
```

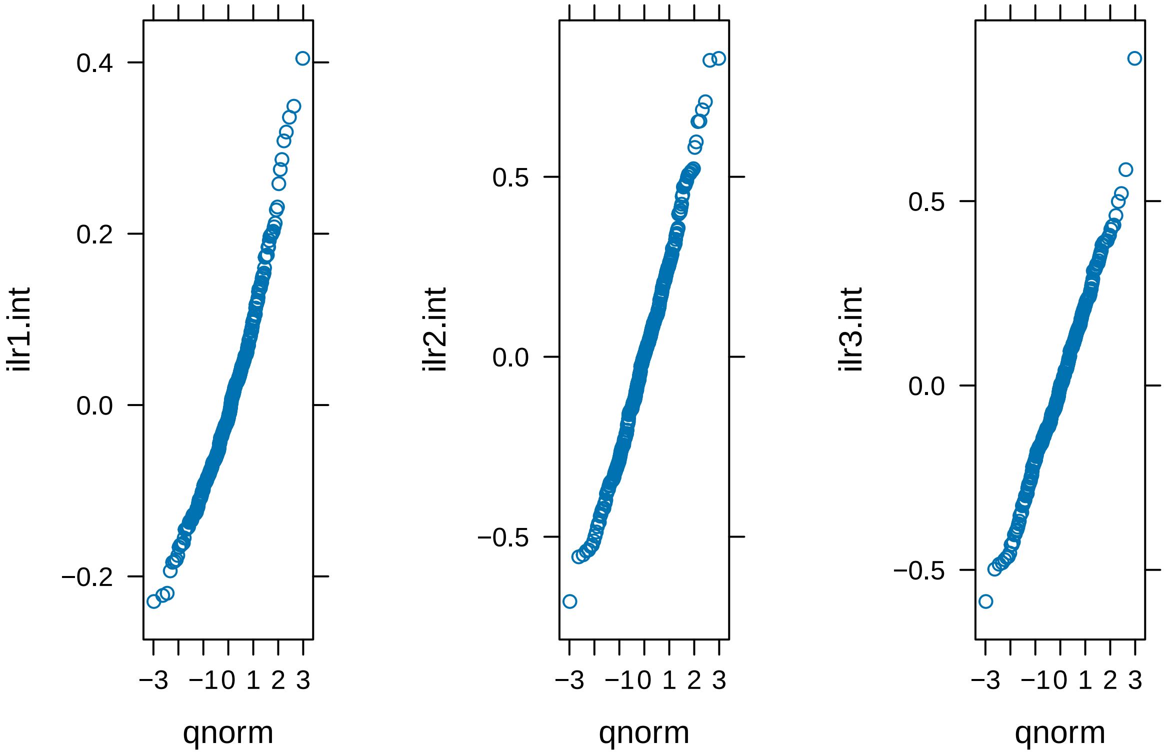


#### Inspecting results

##### Fixed effects

Because we've treated time as a factor var we get a relatively long output. Here we get the intercept for each olr seperately, and then the difference for timepoints 2-5.

```
summary(mod.lme)$tTable
```

```
##                                 Value   Std.Error   DF    t-value      p-value
## ilr.noilr1              0.863602312 0.009909796 3348 87.1463281 0.000000e+00
## ilr.noilr2              1.020861954 0.021471832 3348 47.5442418 0.000000e+00
## ilr.noilr3              0.984494752 0.020033312 3348 49.1428849 0.000000e+00
## ilr.noilr1:timepointT2  0.004643064 0.010952306 3348  0.4239348 6.716406e-01
## ilr.noilr2:timepointT2  0.017258910 0.021122996 3348  0.8170673 4.139481e-01
## ilr.noilr3:timepointT2 -0.013352770 0.021706476 3348 -0.6151515 5.384965e-01
## ilr.noilr1:timepointT3  0.091650045 0.012536943 3348  7.3103982 3.315218e-13
## ilr.noilr2:timepointT3  0.239307905 0.024210459 3348  9.8844846 9.921892e-23
## ilr.noilr3:timepointT3  0.236739644 0.024860594 3348  9.5226866 3.126018e-21
## ilr.noilr1:timepointT4  0.023392000 0.011559207 3348  2.0236682 4.308359e-02
## ilr.noilr2:timepointT4  0.082746221 0.022318826 3348  3.7074629 2.127377e-04
## ilr.noilr3:timepointT4  0.005346990 0.022919132 3348  0.2332981 8.155442e-01
## ilr.noilr1:timepointT5  0.024586447 0.012283896 3348  2.0015187 4.541708e-02
## ilr.noilr2:timepointT5  0.107881370 0.023726738 3348  4.5468269 5.638981e-06
## ilr.noilr3:timepointT5  0.048400999 0.024360281 3348  1.9868818 4.701688e-02
```

##### Random effects

```
#to look at random effects tems
VarCorr(mod.lme)
```

```
## id = pdLogChol(ilr.no - 1)
##              Variance   StdDev    Corr
## ilr.noilr1 0.01481829 0.1217304 ilr.n1 ilr.n2
## ilr.noilr2 0.08894166 0.2982309 0.762
## ilr.noilr3 0.06376742 0.2525221 0.695  0.583
## Residual   0.01720002 0.1311488
```

We can reconstruct matrix as specified in manuscript manually from model output, or can also just extract random effect var/covar matrix with getVarCov() function

```
getVarCov(mod.lme,type = "random.effects")
```

```
## Random effects variance covariance matrix
##              ilr.noilr1 ilr.noilr2 ilr.noilr3
## ilr.noilr1   0.014818   0.027661   0.021373
## ilr.noilr2   0.027661   0.088942   0.043873
## ilr.noilr3   0.021373   0.043873   0.063767
##   Standard Deviations: 0.12173 0.29823 0.25252
```

##### Residual error

This is a little more unweidly. But all the information we need to reconstruct the residual var/covar matrix is in the model output and it can be done easily enough
The residual covariance matrix $E$ can be calulated as:

$$E = DRD$$

where:
- $D$ is a diagonal matrix with standard deviations $\tau_1, \tau_2, \tau_3$.
- $R$ is the correlation matrix:

$$R = \begin{bmatrix} 1 & \rho_{12} & \rho_{13} \\ \rho_{12} & 1 & \rho_{23} \\ \rho_{13} & \rho_{23} & 1 \end{bmatrix}$$

- The resulting $E$ matrix is:

$$E = \begin{bmatrix} \tau_1 & 0 & 0 \\ 0 & \tau_2 & 0 \\ 0 & 0 & \tau_3 \end{bmatrix} \begin{bmatrix} 1 & \rho_{12} & \rho_{13} \\ \rho_{12} & 1 & \rho_{23} \\ \rho_{13} & \rho_{23} & 1 \end{bmatrix} \begin{bmatrix} \tau_1 & 0 & 0 \\ 0 & \tau_2 & 0 \\ 0 & 0 & \tau_3 \end{bmatrix}$$

```
# residual for ilr 1 is just that from model summary
# multiply other ilrs by weighting
tau1 <- sigma(mod.lme)
tau2 <- sigma(mod.lme) * exp(coef(mod.lme$modelStruct$varStruct))[1]
tau3 <- sigma(mod.lme) * exp(coef(mod.lme$modelStruct$varStruct))[2]
```

```r
tau <- c(tau1,tau2,tau3)
D <- diag(tau)
#covariances
rho12 <- as.numeric(coef(mod.lme$modelStruct$corStruct, unconstrained = F)[1])
rho13 <- as.numeric(coef(mod.lme$modelStruct$corStruct, unconstrained = F)[2])
rho23 <- as.numeric(coef(mod.lme$modelStruct$corStruct, unconstrained = F)[3])
R <- matrix(c(
  1,     rho12, rho13,
  rho12, 1,     rho23,
  rho13, rho23, 1
), nrow = 3, byrow = TRUE)

(E <- D %*% R %*% D)
```

```
##             [,1]       [,2]       [,3]
## [1,] 0.01720002 0.02363120 0.02197547
## [2,] 0.02363120 0.06328251 0.04141461
## [3,] 0.02197547 0.04141461 0.06734438
```

### Alternative

Unfortunately, the newer mixed model package lme4 doesn't allow residual side structures, so we can't specify residual var/covar matrix as above (however, G side covariances can be specified as above).

However, this can sometimes be satisfactorily mimicked by adding an additional random effects terms for each observation, as below. Remember, the 'obs' var is simply an indicator variable that groups each set of 3 olr coordinates together.

```r
(mod.lmer <- lme4::lmer(value ~ -1 + ilr.no + ilr.no:timepoint + (ilr.no-1|id) + (ilr.no-1|obs),
                        data = long,
                        control=lmerControl( #need to include lmercontrol arguments
                           check.nobs.vs.nlev="ignore",
                           check.nobs.vs.nRE="ignore",
                           optimizer="bobyqa",
                           calc.derivs = FALSE)))
```

```
## Linear mixed model fit by REML ['lmerMod']
## Formula: value ~ -1 + ilr.no + ilr.no:timepoint + (ilr.no - 1 | id) +
##     (ilr.no - 1 | obs)
##    Data: long
## REML criterion at convergence: -1322.046
## Random effects:
##  Groups   Name        Std.Dev. Corr
##  obs      ilr.noilr1 0.1053
##           ilr.noilr2 0.2391   0.94
##           ilr.noilr3 0.2475   0.84 0.70
##  id       ilr.noilr1 0.1217
##           ilr.noilr2 0.2982   0.76
##           ilr.noilr3 0.2526   0.70 0.58
##  Residual            0.0781
## Number of obs: 3711, groups:  obs, 1237; id, 349
```

```
## Fixed Effects:
##            ilr.noilr1             ilr.noilr2             ilr.noilr3
##              0.863602               1.020862               0.984495
## ilr.noilr1:timepointT2  ilr.noilr2:timepointT2  ilr.noilr3:timepointT2
##              0.004643               0.017259              -0.013352
## ilr.noilr1:timepointT3  ilr.noilr2:timepointT3  ilr.noilr3:timepointT3
##              0.091650               0.239308               0.236740
## ilr.noilr1:timepointT4  ilr.noilr2:timepointT4  ilr.noilr3:timepointT4
##              0.023393               0.082746               0.005348
## ilr.noilr1:timepointT5  ilr.noilr2:timepointT5  ilr.noilr3:timepointT5
##              0.024587               0.107882               0.048403
```

The fixed effects terms should be very similar, however the random effects and residual will be a little different.

```
#for example, random effects from lme
VarCorr(mod.lme)
```

```
## id = pdLogChol(ilr.no - 1)
##            Variance   StdDev    Corr
## ilr.noilr1 0.01481829 0.1217304 ilr.n1 ilr.n2
## ilr.noilr2 0.08894166 0.2982309 0.762
## ilr.noilr3 0.06376742 0.2525221 0.695  0.583
## Residual   0.01720002 0.1311488
```

```
# and then the equivalent lmer mod
VarCorr(mod.lmer)
```

```
##  Groups   Name       Std.Dev. Corr
##  obs      ilr.noilr1 0.105347
##           ilr.noilr2 0.239127 0.938
##           ilr.noilr3 0.247464 0.843 0.700
##  id       ilr.noilr1 0.121737
##           ilr.noilr2 0.298237 0.762
##           ilr.noilr3 0.252557 0.695 0.583
##  Residual            0.078104
```

We can see that the participant level random effects are very similar.

However, the lmer model has an additional random effects output equivalent to the level one variance in the lme model. However, We need to adjust the residual in the lmer model. lmer will provide a residual by default, but this is redundant in this case because we've added an additional random effects term at the observation level where there is only one observation per random effect term. We need to adjust the residual and then we can reconstruct for comparison.

```
# in the lmer model the residual gets confounded in the obs level random effect
# we can adjust after by adding them together
vcv <- (VarCorr(mod.lmer)$obs)
diag(vcv) <- diag(vcv) + sigma(mod.lmer)^2
vcv[1:3,1:3]
```

```
##            ilr.noilr1 ilr.noilr2 ilr.noilr3
```

```
## ilr.noilr1 0.01719823 0.02362996 0.02197263
## ilr.noilr2 0.02362996 0.06328212 0.04141049
## ilr.noilr3 0.02197263 0.04141049 0.06733892
```

Compared to the lme output below.

```
E # from lme output
```

```
##             [,1]       [,2]       [,3]
## [1,] 0.01720002 0.02363120 0.02197547
## [2,] 0.02363120 0.06328251 0.04141461
## [3,] 0.02197547 0.04141461 0.06734438
```

Our level one variance and covariance terms between models are now very similar (after adjusting). The lmer model may be preferred because lme4 is newer and quicker, while also allowing crossed random effects if needed.

### Adding covariates

Extending the model to include time invariant covariates is relatively simple. We simply include an additional fixed effect with an interaction between variables of interest with the dummy variable.

From our base model we'll also include covariates of baseline zBMI and parental education

```
# first, remove participants with missing demographic variables of interest
long <- long %>% dplyr::filter(!if_any(c(Zbmi, Income.cat), is.na))
# now run model
(mod.mrlmm1 <-  nlme::lme(value ~ -1 + ilr.no + ilr.no:timepoint + ilr.no:Zbmi + ilr.no:Income.cat,
                         random = ~ ilr.no - 1 | id, # same as above
                         weights = varIdent(form = ~ 1 | ilr.no), #
                         correlation = corSymm(form = ~1 | id/obs), #
                         control = control,
                         data=long))
```

```
## Linear mixed-effects model fit by REML
##   Data: long
##   Log-restricted-likelihood: 588.5357
##   Fixed: value ~ -1 + ilr.no + ilr.no:timepoint + ilr.no:Zbmi + ilr.no:Income.cat
##                ilr.noilr1                ilr.noilr2                ilr.noilr3
##               0.831244032               0.943657502               0.932839864
##     ilr.noilr1:timepointT2    ilr.noilr2:timepointT2    ilr.noilr3:timepointT2
##               0.008071671               0.011349949              -0.008318101
##     ilr.noilr1:timepointT3    ilr.noilr2:timepointT3    ilr.noilr3:timepointT3
##               0.086365407               0.221222431               0.208394832
##     ilr.noilr1:timepointT4    ilr.noilr2:timepointT4    ilr.noilr3:timepointT4
##               0.025528878               0.092510996               0.009596715
##     ilr.noilr1:timepointT5    ilr.noilr2:timepointT5    ilr.noilr3:timepointT5
##               0.036368406               0.117426367               0.047154846
##           ilr.noilr1:Zbmi           ilr.noilr2:Zbmi           ilr.noilr3:Zbmi
##               0.005228131               0.002833919               0.044555106
## ilr.noilr1:Income.catHigh ilr.noilr2:Income.catHigh ilr.noilr3:Income.catHigh
```

```
##              -0.001166261               0.005412222              -0.068529001
##  ilr.noilr1:Income.catLow  ilr.noilr2:Income.catLow  ilr.noilr3:Income.catLow
##               0.029199820               0.104064958               0.034630885
##
## Random effects:
##  Formula: ~ilr.no - 1 | id
##  Structure: General positive-definite, Log-Cholesky parametrization
##            StdDev    Corr
## ilr.noilr1 0.1114838 ilr.n1 ilr.n2
## ilr.noilr2 0.2861618 0.745
## ilr.noilr3 0.2234271 0.640  0.493
## Residual   0.1183231
##
## Correlation Structure: General
##  Formula: ~1 | id/obs
##  Parameter estimate(s):
##  Correlation:
##   1     2
## 2 0.680
## 3 0.635 0.637
## Variance function:
##  Structure: Different standard deviations per stratum
##  Formula: ~1 | ilr.no
##  Parameter estimates:
##     ilr1     ilr2     ilr3
## 1.000000 2.010929 2.157619
## Number of Observations: 2739
## Number of Groups: 241
```

Random effects and residual as above

```
# random effects matrix as before
getVarCov(mod.mrlmm1,type = "random.effects")
```

```
## Random effects variance covariance matrix
##            ilr.noilr1 ilr.noilr2 ilr.noilr3
## ilr.noilr1   0.012429   0.023781   0.015930
## ilr.noilr2   0.023781   0.081889   0.031534
## ilr.noilr3   0.015930   0.031534   0.049920
##   Standard Deviations: 0.11148 0.28616 0.22343
```

```
# and reconstructing residual as before
tau1 <- sigma(mod.mrlmm1)
tau2 <- sigma(mod.mrlmm1) * exp(coef(mod.mrlmm1$modelStruct$varStruct))[1]
tau3 <- sigma(mod.mrlmm1) * exp(coef(mod.mrlmm1$modelStruct$varStruct))[2]
tau <- c(tau1,tau2,tau3)
D <- diag(tau)
#covariances
rho12 <- as.numeric(coef(mod.mrlmm1$modelStruct$corStruct, unconstrained = F)[1])
rho13 <- as.numeric(coef(mod.mrlmm1$modelStruct$corStruct, unconstrained = F)[2])
rho23 <- as.numeric(coef(mod.mrlmm1$modelStruct$corStruct, unconstrained = F)[3])
R <- matrix(c(
  1,    rho12, rho13,
```

```
  rho12, 1,   rho23,
  rho13, rho23, 1
), nrow = 3, byrow = TRUE)

(E <- D %*% R %*% D)
```

```
##              [,1]       [,2]       [,3]
## [1,] 0.01400036 0.01914775 0.01918559
## [2,] 0.01914775 0.05661515 0.03867239
## [3,] 0.01918559 0.03867239 0.06517611
```

### Manova on fixed effects

This is now relatively simple and results all consistent regardless of SBP used.

```
anova.lme(mod.mrlmm1, type = "marginal")# f
```

```
##                    numDF denDF  F-value p-value
## ilr.no                 3  2475 868.9120  <.0001
## ilr.no:timepoint      12  2475   9.8705  <.0001
## ilr.no:Zbmi            3  2475   4.3998  0.0043
## ilr.no:Income.cat      6  2475   1.8589  0.0842
```

```
car::Anova(mod.mrlmm1, type = 3)# wald
```

```
## Analysis of Deviance Table (Type III tests)
##
## Response: value
##                      Chisq Df Pr(>Chisq)
## ilr.no            2606.736  3  < 2.2e-16 ***
## ilr.no:timepoint   118.446 12  < 2.2e-16 ***
## ilr.no:Zbmi         13.199  3   0.004225 **
## ilr.no:Income.cat   11.153  6   0.083756 .
## ---
## Signif. codes:  0 '***' 0.001 '**' 0.01 '*' 0.05 '.' 0.1 ' ' 1
```

We can see results suggest movement-behvaiour composition changes over timepoints

### Fixed effect coefficient/estimates

```
#ilr coeficients
coef <- tidy(mod.mrlmm1,effect="fixed")%>%
  tidyr::separate(term, into =  c("ilr.no","timepoint"), sep = ":", extra="merge", remove=FALSE)
coef[1:3,4] <- "Intercept"
print(coef[,c(3:6,8:9)],n=24)
```

```
## # A tibble: 24 x 6
##    ilr.no     timepoint      estimate std.error statistic   p.value
##    <chr>      <chr>             <dbl>     <dbl>     <dbl>     <dbl>
##  1 ilr.noilr1 Intercept       0.831     0.0173    48.0    0
##  2 ilr.noilr2 Intercept       0.944     0.0414    22.8    1.91e-104
##  3 ilr.noilr3 Intercept       0.933     0.0357    26.2    2.49e-133
##  4 ilr.noilr1 timepointT2     0.00807   0.0115     0.700  4.84e-  1
##  5 ilr.noilr2 timepointT2     0.0113    0.0233     0.488  6.26e-  1
##  6 ilr.noilr3 timepointT2    -0.00832   0.0249    -0.335  7.38e-  1
##  7 ilr.noilr1 timepointT3     0.0864    0.0129     6.68   2.95e- 11
##  8 ilr.noilr2 timepointT3     0.221     0.0261     8.47   4.09e- 17
##  9 ilr.noilr3 timepointT3     0.208     0.0279     7.48   1.06e- 13
## 10 ilr.noilr1 timepointT4     0.0255    0.0120     2.13   3.32e-  2
## 11 ilr.noilr2 timepointT4     0.0925    0.0242     3.83   1.33e-  4
## 12 ilr.noilr3 timepointT4     0.00960   0.0258     0.372  7.10e-  1
## 13 ilr.noilr1 timepointT5     0.0364    0.0128     2.85   4.41e-  3
## 14 ilr.noilr2 timepointT5     0.117     0.0258     4.56   5.46e-  6
## 15 ilr.noilr3 timepointT5     0.0472    0.0275     1.71   8.67e-  2
## 16 ilr.noilr1 Zbmi            0.00523   0.00698    0.749  4.54e-  1
## 17 ilr.noilr2 Zbmi            0.00283   0.0170     0.166  8.68e-  1
## 18 ilr.noilr3 Zbmi            0.0446    0.0143     3.12   1.82e-  3
## 19 ilr.noilr1 Income.catHigh -0.00117   0.0214    -0.0545 9.57e-  1
## 20 ilr.noilr2 Income.catHigh  0.00541   0.0523     0.103  9.18e-  1
## 21 ilr.noilr3 Income.catHigh -0.0685    0.0437    -1.57   1.17e-  1
## 22 ilr.noilr1 Income.catLow   0.0292    0.0206     1.42   1.55e-  1
## 23 ilr.noilr2 Income.catLow   0.104     0.0503     2.07   3.85e-  2
## 24 ilr.noilr3 Income.catLow   0.0346    0.0420     0.825  4.09e-  1
```

The coefficients above are specific to the basis used. However, we can transform back to compositional coefficients and they will be consistent no matter what SBP used.

Let's just look at coefficients for timepoints as they're most of interest.

```r
comp.coef <- coef[1:15,3:5]
comp.coef <- comp.coef %>% pivot_wider(names_from = ilr.no,
                                       values_from = estimate)
comp.coef <-  as.data.frame(ilrInv(comp.coef[,2:4], V = SBP))
comp.coef$timepoint <-  c("T1","T2","T3","T4","T5")
comp.coef <-  comp.coef[,c(5,1,2,3,4)]
colnames(comp.coef) <- c("Timepoint", "Sleep", "SB", "LPA", "MVPA")
comp.coef[1,2:5] <-  clo(comp.coef[1,2:5], total = 1440)
comp.coef
```

```
##   Timepoint       Sleep          SB         LPA       MVPA
## 1        T1 583.9282509 483.2012872 294.2150662 78.6553957
## 2        T2   0.2517454   0.2517320   0.2468011  0.2497215
## 3        T3   0.2662333   0.2886671   0.2551089  0.1899908
## 4        T4   0.2552904   0.2673232   0.2403129  0.2370735
## 5        T5   0.2574426   0.2716950   0.2432784  0.2275840
```

Here the compositional coefficients can be interpreted as the perturbation vector for each timepoint.
We can compare to the neutral perturbation vector to think about this as reallocations at each timepoint

#### Model based estimates

Alternatively, we can make model-based estimates for each timepoint.

```
# estimated time use as each timepoint from fixed effects
newdata <- expand.grid(ilr.no=c("ilr1","ilr2", "ilr3"),
                       timepoint= c("T1","T2","T3","T4","T5"),
                       Income.cat = "Medium",
                       Zbmi= 0.5107515) #mean bmi
preds <- predict(mod.mrlmm1,newdata=newdata,level = 0,na.action = na.omit)
preds <-  matrix(preds,nrow = 5,ncol = 3,byrow = T)
preds <-  as.data.frame(clo(ilrInv(preds, V = SBP), total = 1440))
preds$Timepoints <- c("T1", "T2", "T3", "T4", "T5")
colnames(preds) <- c("Sleep", "SB", "LPA", "MVPA", "Timepoint")
#values for Figure 1.
preds
```

```
##      Sleep       SB      LPA     MVPA Timepoint
## 1 583.4345 481.8755 297.6392 77.05078        T1
## 2 586.0762 484.0317 293.1147 76.77739        T2
## 3 580.9724 520.2753 283.9990 54.75334        T3
## 4 583.5348 504.6749 280.2254 71.56492        T4
## 5 582.8818 508.0713 280.9971 68.04980        T5
```
