## Supplementary material for "When the outcome is compositional - a method for conducting compositional response linear mixed models for physical activity, sedentary behaviour and sleep research": MRLMM for CoDa model comparison

### MRLMM for CoDa_modelcomparison

```
library(compositions)
library(lme4)
library(forcats)
library(ggplot2); theme_set(theme_bw())
library(plyr)
library(tidyr)
library(dplyr)
library(nlme)
library(broom.mixed)
library(lattice)
library(gridExtra)
```

#### Comparison of CMRLMM to multiple univariate models approach

Compare results for olrs created with two different olr bases using the MRLMM and the multiple univariate models approach.

```
# let's compare model outputs for different SBP when using
# multivariate model vs. 'multiple models' approach
# make copy of data
d2 <- d
d2 <-  d2 %>% dplyr::filter(!if_any(c(Zbmi, Income.cat), is.na)) # remove missing demographics vars
# new ilrs, just use default basis
d2$ilrs <- ilr(d2$a.comp)
d2$ilr1 <- d2$ilrs[,1] #
d2$ilr2 <- d2$ilrs[,2]
d2$ilr3 <- d2$ilrs[,3]
# pivot dataframe
long2 <-  d2 %>% mutate(obs=factor(1:n())) %>%
  pivot_longer(c(ilr1:ilr3), names_to = "ilr.no", values_to = "value")
```

First, make another multivariate model with the new olrs

```
control <-  lmeControl(opt='optim', maxIter = 200, msMaxIter = 200, msMaxEval = 200)
#same model as previously, but with different ilrs
mod.mrlmm2 <-  nlme::lme(value ~ -1 + ilr.no + ilr.no:timepoint + ilr.no:Zbmi + ilr.no:Income.cat,
                        random = ~ ilr.no - 1 | id, # same as above
                        weights = varIdent(form = ~ 1 | ilr.no), #
                        correlation = corSymm(form = ~1 | id/obs), #
                        control = control,
                        data=long2)
```

Now multiple models for olrs specified in manuscript

```
d <-  d %>% dplyr::filter(!if_any(c(Zbmi, Income.cat), is.na)) # remove missing demographics vars
# make 3 seperate models for each ilr and then combine
# first models with first sbp
mod.ilr1 <-  nlme::lme(ilr1 ~ timepoint + Zbmi + Income.cat,
                       random = ~ 1 | id, # same as above
                       control = control,
                       data=d)
mod.ilr2 <-  nlme::lme(ilr2 ~ timepoint + Zbmi + Income.cat,
                       random = ~ 1 | id, # same as above
                       control = control,
                       data=d)
mod.ilr3 <-  nlme::lme(ilr3 ~ timepoint + Zbmi + Income.cat,
                       random = ~ 1 | id, # same as above
                       control = control,
                       data=d)

#basis 2
mod2.ilr1 <-  nlme::lme(ilr1 ~ timepoint + Zbmi + Income.cat,
                        random = ~ 1 | id, # same as above
                        control = control,
                        data=d2)
mod2.ilr2 <-  nlme::lme(ilr2 ~ timepoint + Zbmi + Income.cat,
                        random = ~ 1 | id, # same as above
                        control = control,
                        data=d2)
mod2.ilr3 <-  nlme::lme(ilr3 ~ timepoint + Zbmi + Income.cat,
                        random = ~ 1 | id, # same as above
                        control = control,
                        data=d2)
```

For comparisons sake, the output of the 3 univariate models above can be replicated using a 'stacked' model as well, but this time allowing for heterogeneous variances for each olr, but constraining the off-diagonal elements to be 0. It is easier to do model comparisons with this, so to begin with let's show this model is equivalent.

```
# now for model above but uncorrelated random intercepts
mod.mrlmm1.unrel <-  nlme::lme(value ~ -1 + ilr.no + ilr.no:timepoint + ilr.no:Zbmi + ilr.no:Income.cat
                               random = list(id = pdDiag(form = ~ -1+ factor(ilr.no))),
                               weights = varIdent(form = ~ 1 | factor(ilr.no)),
                               control = control,
                               data=long)
mod.mrlmm1.unrel
```

```
## Linear mixed-effects model fit by REML
##   Data: long
##   Log-restricted-likelihood: 9.028433
##   Fixed: value ~ -1 + ilr.no + ilr.no:timepoint + ilr.no:Zbmi + ilr.no:Income.cat
##               ilr.noilr1                ilr.noilr2                ilr.noilr3
##             0.8311621440              0.9442647526              0.9329646890
##    ilr.noilr1:timepointT2    ilr.noilr2:timepointT2    ilr.noilr3:timepointT2
##             0.0075754914              0.0111342456             -0.0084310686
##    ilr.noilr1:timepointT3    ilr.noilr2:timepointT3    ilr.noilr3:timepointT3
##             0.0862667007              0.2199666576              0.2081600152
```

```
##    ilr.noilr1:timepointT4    ilr.noilr2:timepointT4    ilr.noilr3:timepointT4
##              0.0254292263              0.0921033012              0.0092291692
##    ilr.noilr1:timepointT5    ilr.noilr2:timepointT5    ilr.noilr3:timepointT5
##              0.0361870419              0.1176361024              0.0469527796
##           ilr.noilr1:Zbmi           ilr.noilr2:Zbmi           ilr.noilr3:Zbmi
##              0.0052224673              0.0033782032              0.0445412841
## ilr.noilr1:Income.catHigh ilr.noilr2:Income.catHigh ilr.noilr3:Income.catHigh
##             -0.0008303054              0.0053980773             -0.0684214951
##  ilr.noilr1:Income.catLow  ilr.noilr2:Income.catLow  ilr.noilr3:Income.catLow
##              0.0291595944              0.1020411417              0.0340886599
##
## Random effects:
##  Formula: ~-1 + factor(ilr.no) | id
##  Structure: Diagonal
##         factor(ilr.no)ilr1 factor(ilr.no)ilr2 factor(ilr.no)ilr3  Residual
## StdDev:          0.1111038          0.2850981          0.2232351 0.1184076
##
## Variance function:
##  Structure: Different standard deviations per stratum
##  Formula: ~1 | factor(ilr.no)
##  Parameter estimates:
##     ilr1     ilr2     ilr3
## 1.000000 2.011538 2.156504
## Number of Observations: 2739
## Number of Groups: 241
```

The multivariate unrelated model that assumes no correlation between ilrs in the residual and random effect covariance matrices is now essentially the same as the 3 seperate models approach.

```
#random effects
getVarCov(mod.mrlmm1.unrel,type = "random.effects") # off-diagonal elements constrained to zero
```

```
## Random effects variance covariance matrix
##                    factor(ilr.no)ilr1 factor(ilr.no)ilr2 factor(ilr.no)ilr3
## factor(ilr.no)ilr1           0.012344           0.000000           0.000000
## factor(ilr.no)ilr2           0.000000           0.081281           0.000000
## factor(ilr.no)ilr3           0.000000           0.000000           0.049834
##   Standard Deviations: 0.1111 0.2851 0.22324
```

```
#with each variance of the corresponding univariate models
getVarCov(mod.ilr1,type = "random.effects")
```

```
## Random effects variance covariance matrix
##             (Intercept)
## (Intercept)    0.012344
##   Standard Deviations: 0.1111
```

```
getVarCov(mod.ilr2,type = "random.effects")
```

```
## Random effects variance covariance matrix
##             (Intercept)
## (Intercept)    0.081281
##   Standard Deviations: 0.2851
```

```r
getVarCov(mod.ilr3,type = "random.effects")
```

```
## Random effects variance covariance matrix
##             (Intercept)
## (Intercept)    0.049834
##   Standard Deviations: 0.22324
```

```r
#residual
ilr1var.unc <- as.numeric(VarCorr(mod.mrlmm1.unrel)[4,1])
#multiply other ilrs by weighting
ilr2var.unc <- as.numeric(VarCorr(mod.mrlmm1.unrel)[4,1]) * exp(coef(mod.mrlmm1.unrel$modelStruct$varSt
ilr3var.unc <- as.numeric(VarCorr(mod.mrlmm1.unrel)[4,1]) * exp(coef(mod.mrlmm1.unrel$modelStruct$varSt
# correlations/covariances assumed zero
#reconstruct
res <- format(matrix(c(ilr1var.unc, 0, 0,
                0, ilr2var.unc, 0,
                0, 0, ilr3var.unc),
                nrow = 3,
                ncol = 3,
                byrow = TRUE),scientific = F)
class(res) <- "numeric"
round(res,digits=6)
```

```
##          [,1]    [,2]     [,3]
## [1,] 0.01402 0.00000 0.000000
## [2,] 0.00000 0.05673 0.000000
## [3,] 0.00000 0.00000 0.065202
```

```r
#univariate models
round(as.numeric(VarCorr(mod.ilr1)[2,1]),digits=6)
```

```
## [1] 0.01402
```

```r
round(as.numeric(VarCorr(mod.ilr2)[2,1]),digits=6)
```

```
## [1] 0.05673
```

```r
round(as.numeric(VarCorr(mod.ilr3)[2,1]),digits=6)
```

```
## [1] 0.065202
```

```r
# now some model fit stats
#AIC
AIC(mod.mrlmm1.unrel)
```

```
## [1] 41.94313
```

```r
AIC(mod.ilr1) + AIC(mod.ilr2) + AIC(mod.ilr3)
```

```
## [1] 41.94313
```

```r
# AIC of unrelated mod is the same as sum of univariate mods

#loglik
logLik(mod.mrlmm1.unrel)
```

```
## 'log Lik.' 9.028433 (df=30)
```

```r
logLik(mod.ilr1) + logLik(mod.ilr2) + logLik(mod.ilr3)
```

```
## 'log Lik.' 9.028433 (df=10)
```

```r
# loglikelihood of unrelated mod is the same as sum of univariate mods

# deviance
deviance(update(mod.mrlmm1.unrel,method="ML"))
```

```
## 'log Lik.' -163.5685 (df=30)
```

```r
deviance(update(mod.ilr1,method="ML")) + deviance(update(mod.ilr2,method="ML")) + deviance(update(mod.il
```

```
## 'log Lik.' -163.5685 (df=10)
```

```r
# deviance of unrelated mod is the same as sum of univariate mods

# and now BLUP estimates, predicted values, residuals etc.
# random effects
round(head(ranef(mod.mrlmm1.unrel)),digits = 4) # multivariate unrelated model
```

```
##         factor(ilr.no)ilr1 factor(ilr.no)ilr2 factor(ilr.no)ilr3
## LOH002C            -0.0545            -0.0131             0.2370
## LOH004C            -0.1181            -0.4689            -0.3386
## LOH005C             0.0215             0.1029             0.1077
## LOH006C             0.2063             0.5181             0.0973
## LOH008C             0.2052             0.5036             0.3866
## LOH009C             0.0572             0.1619            -0.0153
```

```r
round(head(bind_cols(ranef(mod.ilr1),ranef(mod.ilr2),ranef(mod.ilr3))),digits = 4) # univariate models
```

```
##         (Intercept)...1 (Intercept)...2 (Intercept)...3
## LOH002C         -0.0545         -0.0131          0.2370
## LOH004C         -0.1181         -0.4689         -0.3386
## LOH005C          0.0215          0.1029          0.1077
## LOH006C          0.2063          0.5181          0.0973
## LOH008C          0.2052          0.5036          0.3866
## LOH009C          0.0572          0.1619         -0.0153
```

```r
#residuals
round(head(as.data.frame(matrix(resid(mod.mrlmm1.unrel), # multivariate unrelated model
                                                  ncol = 3,
                                                  byrow = T))),digits = 4)
```

```
##        V1      V2      V3
## 1 -0.0346 -0.1256  0.0045
## 2 -0.0526  0.1290 -0.1803
## 3  0.0641  0.1134 -0.0390
## 4 -0.0634  0.0046 -0.1248
## 5  0.0791  0.0932 -0.0154
## 6  0.0386 -0.0983 -0.0579
```

```r
round(head(bind_cols(resid(mod.ilr1),resid(mod.ilr2),resid(mod.ilr3))),digits = 4) # univariate
```

```
## # A tibble: 6 x 3
##       ...1    ...2    ...3
##      <dbl>   <dbl>   <dbl>
## 1 -0.0346 -0.126   0.0045
## 2 -0.0526  0.129  -0.180
## 3  0.0641  0.113  -0.039
## 4 -0.0634  0.0046 -0.125
## 5  0.0791  0.0932 -0.0154
## 6  0.0386 -0.0983 -0.0579
```

```r
#fitted values at highest level
round(head(as.data.frame(matrix(predict(mod.mrlmm1.unrel), # multivariate unrelated model
                    ncol = 3,
                    byrow = T))),digits = 4)
```

```
##       V1     V2     V3
## 1 0.7842 0.9419 1.1726
## 2 0.7431 0.5779 0.6358
## 3 0.8872 1.1527 1.1205
## 4 1.0392 1.4635 1.0456
## 5 1.0645 1.5493 1.3448
## 6 0.8806 1.1012 0.8516
```

```r
#
round(head(as.data.frame(matrix(c(predict(mod.ilr1,level = 1),
                      predict(mod.ilr2,level = 1),
                      predict(mod.ilr3,level = 1)), #univariate
                    ncol = 3,
                    byrow = F))),digits = 4)
```

```
##       V1     V2     V3
## 1 0.7842 0.9419 1.1726
## 2 0.7431 0.5779 0.6358
## 3 0.8872 1.1527 1.1205
## 4 1.0392 1.4635 1.0456
## 5 1.0644 1.5493 1.3448
## 6 0.8806 1.1012 0.8516
```

And multiple models with other olr base

```
# now for model above but uncorrelated random intercepts and errors
mod.mrlmm2.unrel <-  nlme::lme(value ~ -1 + ilr.no + ilr.no:timepoint + ilr.no:Zbmi + ilr.no:Income.cat
                            random = list(id = pdDiag(form = ~ -1+ factor(ilr.no))),
                            weights = varIdent(form = ~ 1 | factor(ilr.no)),
                            control = lmeControl(
                              opt='optim', maxIter = 200, msMaxIter = 200, msMaxEval = 200),
                            data=long2)
```

#### log-likelihood

Now that we have our models, we can compare them.

```
# update to ml for model comparison
mod.mrlmm1 <- update(mod.mrlmm1, method = "ML")
mod.mrlmm2 <- update(mod.mrlmm2, method = "ML")
mod.mrlmm1.unrel <- update(mod.mrlmm1.unrel, method = "ML")
mod.mrlmm2.unrel <- update(mod.mrlmm2.unrel, method = "ML")
# compare mods. .c suffix for correlated models and .u for uncorrelated (multiple models approach)
loglik.c <- c(logLik(mod.mrlmm1),logLik(mod.mrlmm2))
names(loglik.c) <- c("MRLMM1", "MRLMM2")
loglik.u <- c(logLik(mod.mrlmm1.unrel),logLik(mod.mrlmm2.unrel))
names(loglik.u) <- c("UnrelMod1", "UnrelMod2")

loglik.c #same
```

```
##   MRLMM1   MRLMM2
## 666.4208 666.4208
```

```
loglik.u # different
```

```
## UnrelMod1 UnrelMod2
##  81.78424 384.25470
```

log-likelihood of multivaraite models is the same, not so for unrelated models

#### AIC

```
AIC.c <- c(AIC(mod.mrlmm1),AIC(mod.mrlmm2))
names(AIC.c) <- c("MRLMM1", "MRLMM2")
AIC.u <- c(AIC(mod.mrlmm1.unrel),AIC(mod.mrlmm2.unrel))
names(AIC.u) <- c("UnrelMod1", "UnrelMod2")
AIC.c # mrlmm
```

```
##    MRLMM1    MRLMM2
## -1260.842 -1260.842
```

```r
AIC.u # uncorrelated mods
```

```
## UnrelMod1 UnrelMod2
## -103.5685 -708.5094
```

same true for AIC

#### BIC

```r
BIC.c <- c(BIC(mod.mrlmm1),BIC(mod.mrlmm2))
names(BIC.c) <- c("MRLMM1", "MRLMM1")
BIC.u <- c(BIC(mod.mrlmm1.unrel),BIC(mod.mrlmm2.unrel))
names(BIC.u) <- c("UnrelMod1", "UnrelMod2")
BIC.c # mrlmm
```

```
##    MRLMM1    MRLMM1
## -1047.889 -1047.889
```

```r
BIC.u # uncorrelated mods
```

```
##  UnrelMod1  UnrelMod2
##   73.89196 -531.04896
```

and BIC
#### deviance

```r
deviance.c <- c(deviance(mod.mrlmm1),deviance(mod.mrlmm2))
names(deviance.c) <- c("MRLMM1", "MRLMM1")
deviance.u <- c(deviance(mod.mrlmm1.unrel),deviance(mod.mrlmm2.unrel))
names(deviance.u) <- c("UnrelMod1", "UnrelMod2")
deviance.c # mrlmm
```

```
##    MRLMM1    MRLMM1
## -1332.842 -1332.842
```

```r
deviance.u # uncorrelated mods
```

```
## UnrelMod1 UnrelMod2
## -163.5685 -768.5094
```

#### Multivariate F test

```r
## f-test on fixed effects
anova.lme(mod.mrlmm1,type="marginal")
```

```
##                    numDF denDF  F-value p-value
## ilr.no                  3  2475 875.7943  <.0001
## ilr.no:timepoint       12  2475   9.8446  <.0001
## ilr.no:Zbmi             3  2475   4.4338  0.0041
## ilr.no:Income.cat       6  2475   1.8756  0.0813
```

```r
anova.lme(mod.mrlmm2,type="marginal")
```

```
##                    numDF denDF  F-value p-value
## ilr.no                  3  2475 875.8032  <.0001
## ilr.no:timepoint       12  2475   9.8446  <.0001
## ilr.no:Zbmi             3  2475   4.4340  0.0041
## ilr.no:Income.cat       6  2475   1.8755  0.0813
```

```r
anova.lme(mod.mrlmm1.unrel,type="marginal")
```

```
##                    numDF denDF   F-value p-value
## ilr.no                  3  2475 1180.8785  <.0001
## ilr.no:timepoint       12  2475   17.7869  <.0001
## ilr.no:Zbmi             3  2475    3.4743  0.0154
## ilr.no:Income.cat       6  2475    2.5843  0.0169
```

```r
anova.lme(mod.mrlmm2.unrel,type="marginal")
```

```
##                    numDF denDF  F-value p-value
## ilr.no                  3  2475 455.0358  <.0001
## ilr.no:timepoint       12  2475  13.8847  <.0001
## ilr.no:Zbmi             3  2475   2.6860  0.0451
## ilr.no:Income.cat       6  2475   2.3800  0.0270
```

By constraining the off-diagonal elements of the G and E covariance matrices to be equal to zero, the models constructed with olr coordinates constructed with a different basis are not the same. This is because the assumption that olr coordinates are uncorrelated is generally untenable (though this may not always be the case). However, (correlated) multivariate response models are equivalent, regardless of the olr basis chosen.
