## Supplementary material for "When the outcome is compositional - a method for conducting compositional response linear mixed models for physical activity, sedentary behaviour and sleep research": Mathematical background

Let

$$\mathbf{z}_{ij} = \boldsymbol{\beta}_0 + \boldsymbol{\beta}_1 t_{ij} + \mathbf{b}_{0i} + \boldsymbol{\varepsilon}_{ij}$$

be the CMRLMM where $\mathbf{z}_{ij} = [z_{1ij}, z_{2ij}, \dots, z_{(D-1)ij}]$ are the olr-coordinates of the composition and $\boldsymbol{\beta}_0 = [\beta_{01}, \beta_{02}, \dots, \beta_{0(D-1)}]$, $\boldsymbol{\beta}_1 = [\beta_{11}, \beta_{12}, \dots, \beta_{1(D-1)}]$ the vectors of coefficients. The random effect and error are assumed normally distributed:

$$\mathbf{b}_{0i} = \begin{pmatrix} b_{01i} \\ b_{02i} \\ \vdots \\ b_{0(D-1)i} \end{pmatrix} \sim \mathcal{MVN}(\mathbf{0}_{\boldsymbol{D-1}}, \mathbf{G})$$

$$\boldsymbol{\varepsilon}_{ij} = \begin{pmatrix} \varepsilon_{1ij} \\ \varepsilon_{2ij} \\ \vdots \\ \varepsilon_{(D-1)ij} \end{pmatrix} \sim \mathcal{MVN}(\mathbf{0}_{\boldsymbol{D-1}}, \mathbf{E})$$

where the covariance matrices respectively are

$$\mathbf{G} = \left[\rho_{ij}^{(g)} \sigma_i \sigma_j\right], \qquad i, j = 1,2, \dots, D-1$$

and

$$\mathbf{E} = \left[\rho_{ij}^{(e)} \tau_i \tau_j\right], \qquad i, j = 1,2, \dots, D-1$$

Let $\boldsymbol{\Phi}$ be a change of olr-basis, where $\boldsymbol{\Phi} \cdot \boldsymbol{\Phi}^{\boldsymbol{T}} = \boldsymbol{\Phi}^{\boldsymbol{T}} \cdot \boldsymbol{\Phi} = \mathbf{Id}_{\boldsymbol{D-1}}$. Note that $\boldsymbol{\Phi}$ can be interpreted a “rotation” matrix.

When a change of basis is applied, we obtain new olr-coordinates $\mathbf{z}_{ij}^* = \boldsymbol{\Phi} \cdot \mathbf{z}_{ij}$ and the CMRLMM becomes

$$\mathbf{z}_{ij}^* = \boldsymbol{\Phi} \cdot \mathbf{z}_{ij} = \boldsymbol{\Phi} \cdot \boldsymbol{\beta}_0 + \boldsymbol{\Phi} \cdot \boldsymbol{\beta}_1 t_{ij} + \boldsymbol{\Phi} \cdot \mathbf{b}_{0i} + \boldsymbol{\Phi} \cdot \boldsymbol{\varepsilon}_{ij} = \boldsymbol{\beta}_0^* + \boldsymbol{\beta}_1^* t_{ij} + \mathbf{b}_{0i}^* + \boldsymbol{\varepsilon}_{ij}^*,$$

where

$$\mathbf{b}_{0i}^* \sim \mathcal{MVN}(\mathbf{0}_{\boldsymbol{D-1}}, \boldsymbol{\Phi} \cdot \mathbf{G} \cdot \boldsymbol{\Phi}^{\boldsymbol{T}}) = \mathcal{MVN}(\mathbf{0}_{\boldsymbol{D-1}}, \mathbf{G}^*)$$

$$\boldsymbol{\varepsilon}_{ij}^* \sim \mathcal{MVN}(\mathbf{0}_{\boldsymbol{D-1}}, \boldsymbol{\Phi} \cdot \mathbf{E} \cdot \boldsymbol{\Phi}^{\boldsymbol{T}}) = \mathcal{MVN}(\mathbf{0}_{\boldsymbol{D-1}}, \mathbf{E}^*).$$

We consider three different cases:

1. “Spherical”: if $\mathbf{G} = \sigma \cdot \mathbf{Id}_{\boldsymbol{D-1}}$ and $\mathbf{E} = \tau \cdot \mathbf{Id}_{\boldsymbol{D-1}}$ then $\mathbf{G}^* = \mathbf{G}$ and $\mathbf{E}^* = \mathbf{E}$. Therefore, when using multiple-univariate approach or multivariant approach, the results are the same. In addition, in both approaches the results expressed in terms of raw compositions are invariant under a change of basis.

2. “Ellipses with axes that are parallel to coordinate axes”: in this case, the matrices are $\mathbf{G} = \begin{bmatrix} \sigma_1^2 & 0 & \cdots & 0 \\ 0 & \sigma_2^2 & \cdots & 0 \\ 0 & 0 & \ddots & \vdots \\ 0 & 0 & 0 & \sigma_{(D-1)}^2 \end{bmatrix}$ and $\mathbf{E} = \begin{bmatrix} \tau_1^2 & 0 & \cdots & 0 \\ 0 & \tau_2^2 & \cdots & 0 \\ 0 & 0 & \ddots & \vdots \\ 0 & 0 & 0 & \tau_{(D-1)}^2 \end{bmatrix}$, that is, the matrices are diagonal, being $\rho_{ij}^{(g)} = \rho_{ij}^{(e)} = 0$, for $i < j = 1,2, \dots, D-1$. Again, when using multiple-univariate approach or multivariant approach, the results are the same. However, when a change of olr-basis is applied the “ellipses” are rotated and they no longer have axes parallel to the coordinate axes. That is, $\mathbf{G}^* = \mathbf{\Phi} \cdot \mathbf{G} \cdot \mathbf{\Phi}^{\boldsymbol{T}}$ and $\mathbf{E}^* = \mathbf{\Phi} \cdot \mathbf{E} \cdot \mathbf{\Phi}^{\boldsymbol{T}}$, are not diagonal matrices. Therefore, the results are different when using multiple-univariate approach and multivariant approach to the new olr-coordinates $\mathbf{z}_{ij}^*$. Importantly, only the multivariate method does verify the property of invariance under a change of basis.
3. “Ellipses with axes that are not parallel to coordinate axes”: in this general case, the matrices are

$$\mathbf{G} = \begin{bmatrix} \sigma_1^2 & \rho_{12}^{(g)}\sigma_1\sigma_2 & \cdots & \rho_{1(D-1)}^{(g)}\sigma_1\sigma_{(D-1)} \\ \square & \sigma_2^2 & \cdots & \rho_{2(D-1)}^{(g)}\sigma_2\sigma_{(D-1)} \\ \square & \square & \ddots & \vdots \\ \square & \square & \square & \sigma_{(D-1)}^2 \end{bmatrix} \text{and}$$

$$\mathbf{E} = \begin{bmatrix} \tau_1^2 & \rho_{12}^{(e)}\tau_1\tau_2 & \cdots & \rho_{1(D-1)}^{(e)}\tau_1\tau_{(D-1)} \\ \square & \tau_2^2 & \cdots & \rho_{2(D-1)}^{(e)}\tau_2\tau_{(D-1)} \\ \square & \square & \ddots & \vdots \\ \square & \square & \square & \tau_{(D-1)}^2 \end{bmatrix},$$

   that is, the matrices are not diagonal. In this case, the results are different when using multiple-univariate approach and multivariate approach, because the first approach assumes that $\rho_{ij}^{(g)} = \rho_{ij}^{(e)} = 0$, for $i < j = 1,2, \dots, D-1$ , which is not the case. In addition, again, only the multivariate method does verify the property of invariance under a change of basis.

In conclusion, the multiple-univariate approach only is adequate for the case of both random effect and error are assumed “spherically” normally distributed (case 1).
